# Subnational equity in the delivery of primary health care interventions during health shocks: lessons learned from an implementation research study in Rwanda

**DOI:** 10.1101/2025.03.28.25324830

**Authors:** Alemayehu Amberbir, Felix Sayinzoga, Aline Uwimana, Amelia VanderZanden, Jovial Thomas Ntawukuriryayo, Ernest Tambo, Anil Krishna, Lisa R Hirschhorn, Agnes Binagwaho

## Abstract

The COVID-19 pandemic has brought about significant disruptions to health care delivery worldwide, including in Rwanda. Countries experienced variable disruptions both at the national and the subnational level. Here we report results from mixed methods implementation research on lessons learned from Rwanda to mitigate inequity in the delivery of health care interventions during health shocks as in the COVID-19 pandemic. To estimate the coverage of primary health care interventions known to reduce under-5 mortality during the initial period of COVID-19 in Rwanda, we analyzed existing data from the health management information system. Using the available administrative health management information system data in Rwanda during 2019 and 2020, we calculated i) cumulative and ii) monthly disruption ratios of number of cases of facility-based delivery, number of four or more antenatal care visits, and number of diarrheal cases treated at health facility and community levels in 2019 and in 2020. We conducted key informant interviews between February to April 2021 with policymakers, donors, implementing partners, and direct health services providers to identify barriers and facilitators of subnational variability in the delivery of primary health care interventions, as well as implementation strategies, across Rwanda’s districts. We report district-level results of cumulative and monthly disruptions for three interventions. We found minimal disruption across most districts in Rwanda in the first phase of the COVID-19 pandemic (March to December 2020) and, furthermore, we found minimal subnational variability across districts. Implementation strategies such as community health worker interventions, community engagement and education, provision of transport, and command posts, were important in ensuring minimal disruption across most districts. Rwanda’s focus on equity likely helped to strengthen facilitating contextual factors including a culture of accountability and a strong pre-existing community health system and structure, which contributed to the low level of disruption and minimal subnational variability in the interventions studied. Rwanda’s experience offers potentially transferable knowledge for policymakers and decision-makers in other regions and countries to minimize disruptions at the subnational and national levels to essential health services during future health shocks.

**Key messages:**

- In the first phase of the COVID-19 pandemic, we found minimal disruption across most districts in Rwanda, similar to previous findings of minimal disruption for the country as a whole and, furthermore, we found minimal subnational variability in disruption across districts.
- We found that implementation strategies such as community health worker interventions, community engagement and education, provision of transport, and command posts (the National Joint Task Force composed of multidisciplinary teams and supported by subnational task force from all 30 districts to ensure all the coordination around COVID-19 runs smoothly and supports health care delivery) were important factors in delivering minimal disruption across most districts in Rwanda during the first period of COVID-19.
- Rwanda’s focus on equity likely helped to strengthen facilitating contextual factors including a culture of accountability and a strong pre-existing community health system and structure, which contributed to the low level of disruption and minimal subnational variability in the interventions we considered.
- The capacity to conduct both national and subnational analysis of disruption and understand strategies which were applied is valuable for providing meaningful lessons learned for communities in other regions and countries.

## Introduction

The COVID-19 pandemic has brought about significant disruptions to health care delivery worldwide, including in Rwanda.[1,2] Countries experienced variable disruptions both at the national and the subnational levels. Subnational disruption of health care delivery due to COVID-19 refers to challenges faced at regional or district levels in providing essential health services while combating the spread of the virus. In Rwanda, as in many other countries, the immediate response to the pandemic necessitated a redirection of resources and focus towards COVID-19 prevention and treatment.[3] This shift in priorities, coupled with measures such as lockdowns, travel restrictions, and social distancing protocols, has had a profound impact on the delivery of non-COVID-19 related health care services.

The first case of COVID-19 was reported in Rwanda on March 14, 2020 and immediate intervention measures including contact tracing and complete national lockdowns were put in place. Between March to December 2020, 10 months after its first case, Rwanda recorded 10,316 cases and 133 COVID-19-related deaths with a case fatality rate of 1.3%.[4] One week after the first case, the government extended the complete national lockdown to the 30^th^ of April 2020 to minimize the virus spreading across the country, with preservation of essential services such as health care and food-related businesses.[5,6] After the complete lockdown lifted, additional partial lockdowns were applied in different districts at different times depending on the number of cases (Figure 1).[7] Throughout the pandemic, Rwanda implemented all the public health measures known to reduce the spread of the pandemic. This included testing and contact tracing, isolation and quarantine, mask wearing, prohibition of mass gatherings, social distancing, temperature checks at airports and public buildings, the creation of separate COVID-19 treatment centers, and vaccination.[8]

**Figure 1.**
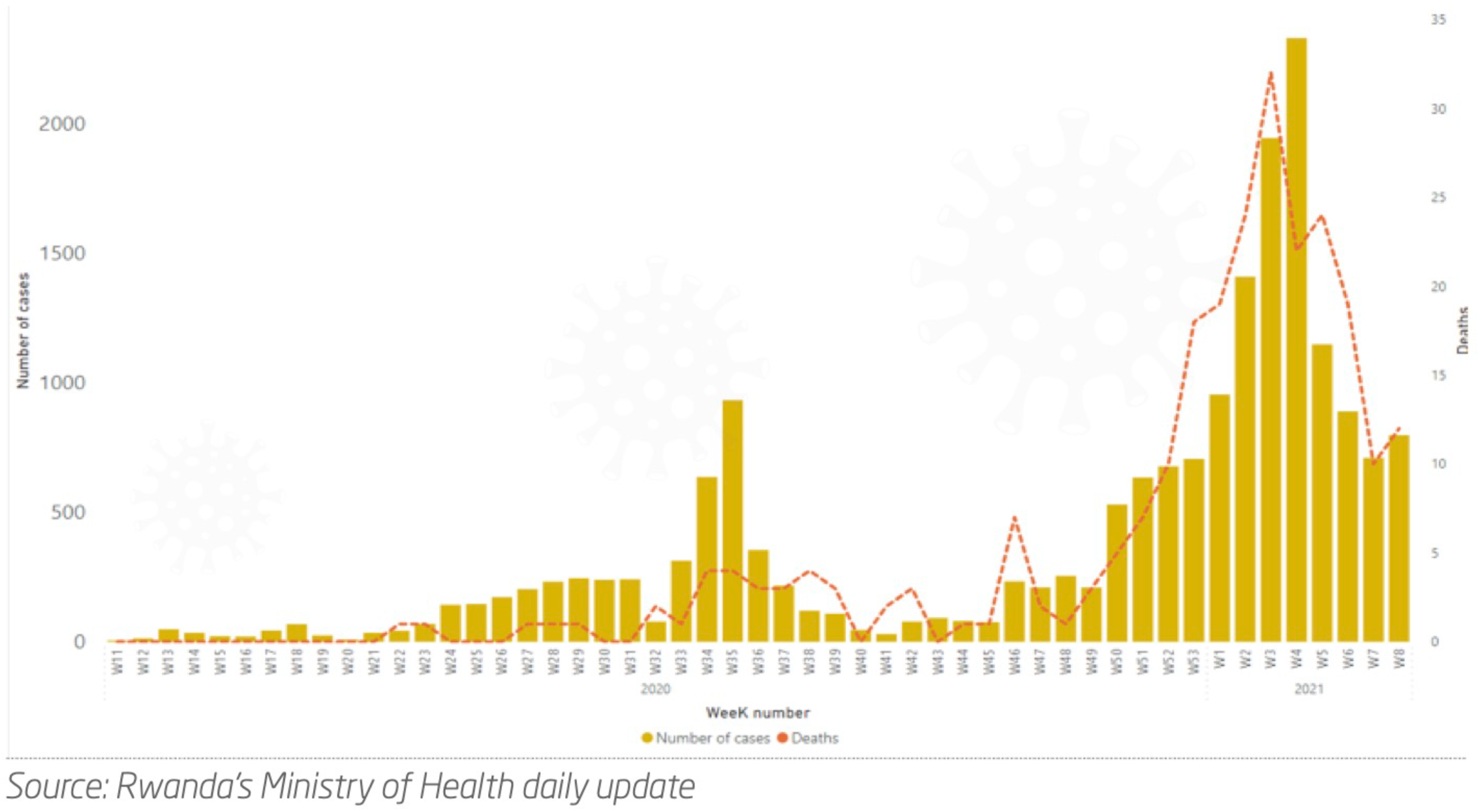
Cases and deaths from COVID-19 in Rwanda, March 2020 to February 2021. *Source*: *Rwanda Biomedical Center Weekly Epidemiological Bulletin*.

One of the key challenges of COVID-19 prevention and response measures, beyond the disruption due to lockdowns, has been the resulting strain placed on health care facilities and systems. The increased demand for COVID-19 care has stretched resources and capacities, potentially resulting in reduced access and availability of other critical health services – and impacting efforts towards the Sustainable Development Goals targeting health.[1,9] Additionally, fear and misinformation surrounding the virus may have discouraged individuals from seeking routine medical care or accessing necessary treatments.[2] These disruptions are particularly felt at the subnational level, where health care infrastructure and resources may already be limited. This includes difficulties in providing essential health services such as immunizations, prenatal care, chronic disease management, and routine screenings. Disparate communities may also experience disparities in access to COVID-19 testing and treatment, and preexisting disparities in access and resources, exacerbating existing health inequities.[2] By navigating these hurdles and adapting to the evolving situation, Rwanda sought to minimize the subnational disruption of health care delivery caused by COVID-19 including maintaining one of the lowest case fatality rates in Africa.[4]

As part of a larger project to explore if and how health system-delivered evidence-based interventions targeting amenable under-5 mortality (U5M) were maintained during the early period of COVID-19, we conducted this study to understand the cumulative and monthly disruption as well as subnational variability of the delivery of primary health care interventions including facility-based delivery, four or more ANC visits (ANC4+), and number of diarrheal cases reported at the facility and community during the period of COVID-19. We reported the lessons learned from Rwanda to mitigate inequity in the delivery of health care interventions during health shocks as in COVID-19.

## Methods

### Study design

The study utilized a mixed methods implementation research approach. The interventions known to reduce U5M we examined included facility-based delivery, ANC4+ visits, and the treatment of diarrheal cases in the community and health facilities in Rwanda and subnationally by district. These interventions were selected based on their relevance and the complex skills required to save lives of children under 5 during critical moments. The work was guided by our implementation research framework designed as part of earlier work to understand how some countries reduced U5M faster than their regional or economic peers (Figure 2).[10] We utilized this framework to understand strategies and contextual factors subnationally, at facilities, and across the interventions implemented for the reduction of U5M during the COVID-19 pandemic period.

**Figure 2.**
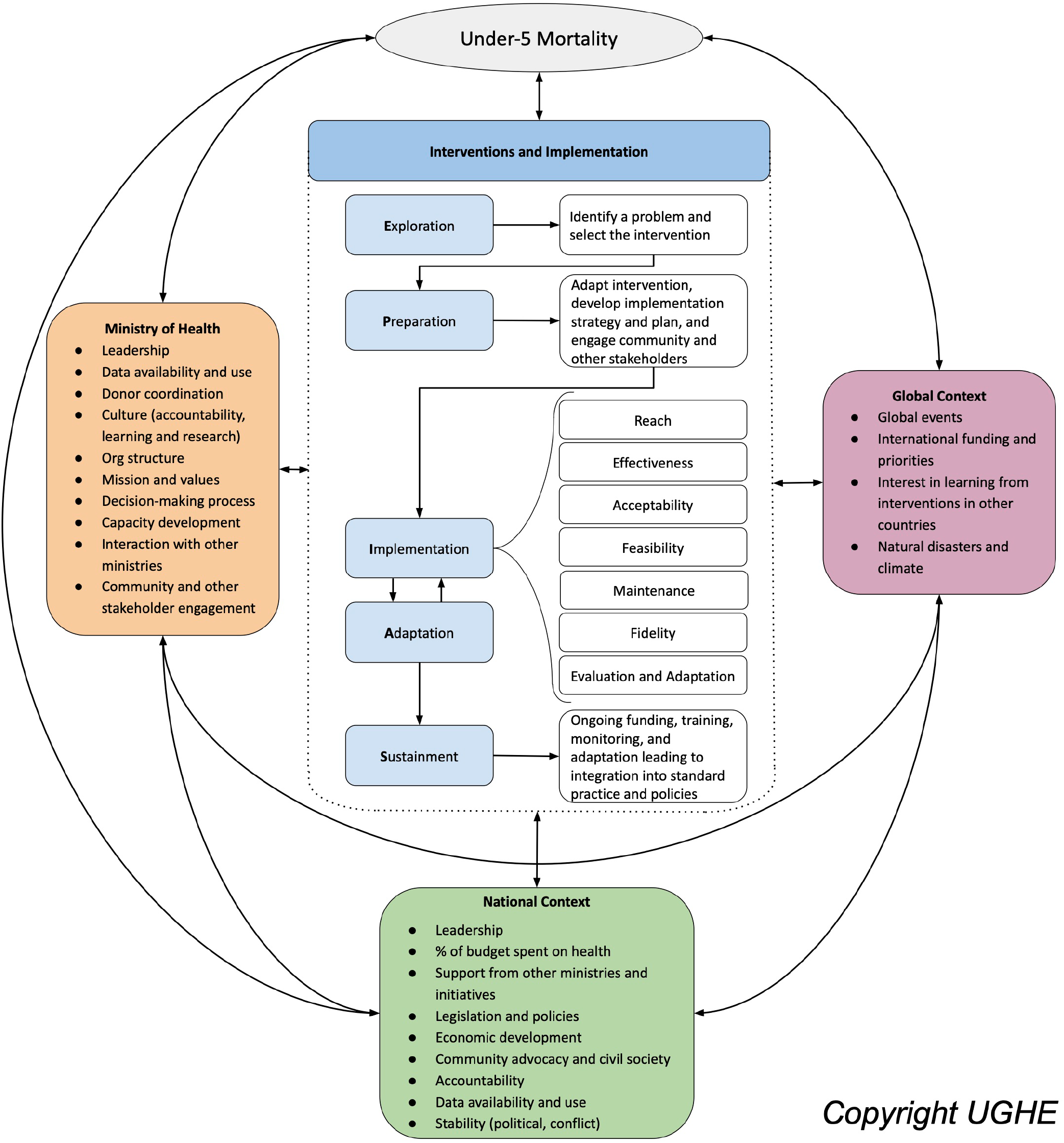
Implementation research framework for understanding evidence-based interventions to reduce under-5 mortality[10]

### Data collection and measures

To estimate the disruption to the targeted interventions known to reduce U5M before and during the initial period of COVID-19 in Rwanda, we analyzed existing data from the health management information system (HMIS). We first collated total monthly facility-based delivery, ANC4+, and number of diarrheal cases treated in health facilities and in the community between January to December in 2019 (prior the period of COVID-19) and between March to December 2020 (during the period of COVID-19), as reported by the Ministry of Health in Rwanda. We conducted selected key informant interviews between February to April 2021 with policymakers, donors, implementing partners, and direct health services providers to identify barriers and facilitators of subnational variability as well as relevant implementation strategies in the delivery of primary health care interventions across Rwanda’s districts.

### Data analysis

Using the available administrative HMIS data in Rwanda during 2019 and 2020, we calculated i) cumulative and ii) monthly disruption ratios of number of facility-based deliveries, number of ANC4+ visits, and number of diarrheal cases treated at health facility and community level in 2019 and in 2020. We adopted a similar methodology as reported by Causey and colleagues.[1]

The cumulative disruption ratio is the disruption ratio, relative to the change in number of cases reported (for example number of facility-based deliveries reported) from 2019 to 2020, as measured by the ratio between reported cases in January and February of 2020 versus the same months in 2019. This approach uses January and February of 2019 as a baseline.

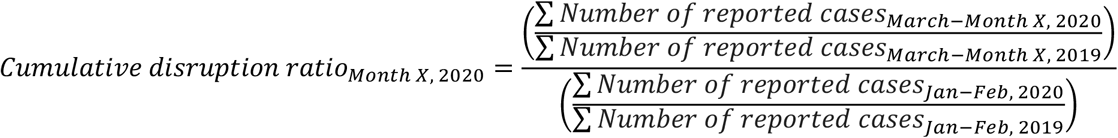

The monthly disruption ratio is the ratio of the number of cases reported – for example, the number of reported facility-based deliveries – during a given month in 2020 and the corresponding month in 2019 using the formula below. The monthly data for 2019 and 2020 were obtained from administrative data available in HMIS.

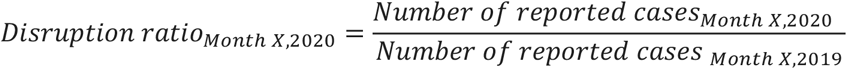

A disruption ratio (cumulative or monthly) of 0 would indicate complete disruption of service delivery (e.g. facility-based delivery, number of ANC4+ visits, and number of cases of diarrheal diseases as proxy for access to primary health care services) between March 2020 (when the first COVID-19 cases were reported in Rwanda), and the month in question, whereas a ratio of 1 would indicate that the expected number of service delivery had been delivered during that period. A disruption ratio above 1 indicates that more than the expected number of services had been delivered for the respective intervention.

The qualitative data from key informant interviews with key health system policymakers, decision-makers, and implementors from national to primary health center levels were audio recorded, transcribed, and analyzed using a thematic framework.[11] We used explanatory mixed methods to understand the patterns of monthly disruption ratio nationally and by district, through the themes which emerged from the key informant interviews.

### Ethics

Our study was approved by the Rwandan National Ethics Committee (Approval No: 1121/RNEC/2020), Rwandan National Health Research Committee (Approval No: NHRC/2021/PROT/014) and the Rwandan MOH (Approval No: 20/805/MIN/2021). We obtained written informed consent from individual key informants and anonymity and confidentiality were maintained throughout the project.

## Results

### Facility-based deliveries

Cumulative disruption: In most districts, there were minimal cumulative disruptions to facility-based deliveries from March to December 2020, with the disruption ratio at 1 (Figure 3). Gasabo, Kirehe, Kayonza, Ngoma, Nyabihu, Nyamagabe, Nyarugenge, Rulindo, and Rutsiro had lower than expected facility-based delivery services (disruption ratio <1), though in most cases the disruptions were small. An exception was Rusizi which had a disruption ratio above 1, with more than expected facility-based deliveries from March to December 2020.

**Figure 3.**
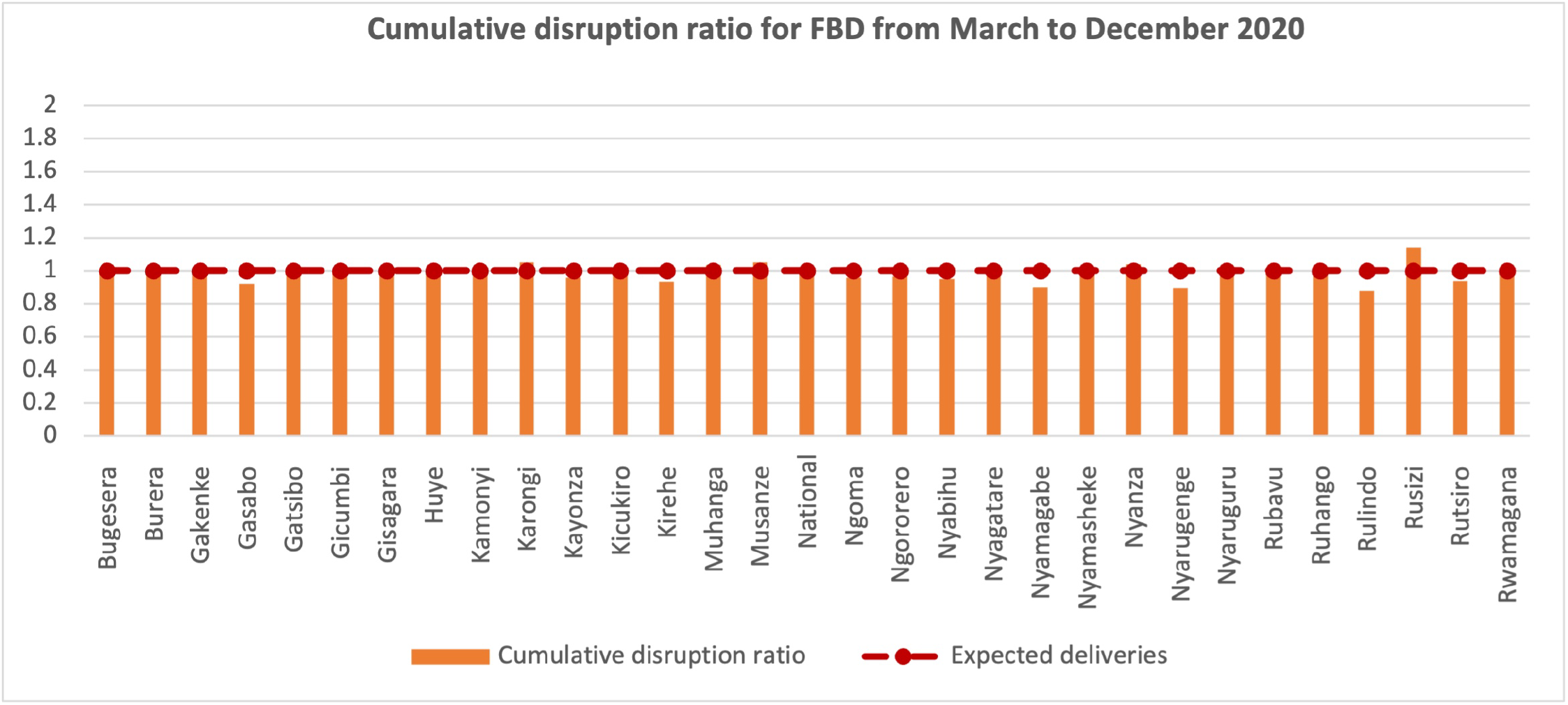
Monthly cumulative disruption ratios of facility-based delivery in Rwanda between March-December 2020

Monthly disruption: For monthly disruption (Figure 4) four districts had higher than expected facility-based deliveries, although most were only 1-2 months. Muhanga had higher numbers of facility-based deliveries in May and September 2020 than expected. Musanze had higher numbers of facility-based deliveries in May and November 2020 than expected. Nyanza had higher numbers of facility-based deliveries in September 2020 than expected. Rusizi had higher numbers of deliveries from April to December 2020 than expected. Figure 4 shows monthly disruption nationally and for one district in each of Rwanda’s five provinces for illustration; full district-level results are available in the supplemental materials.

**Figure 4.**
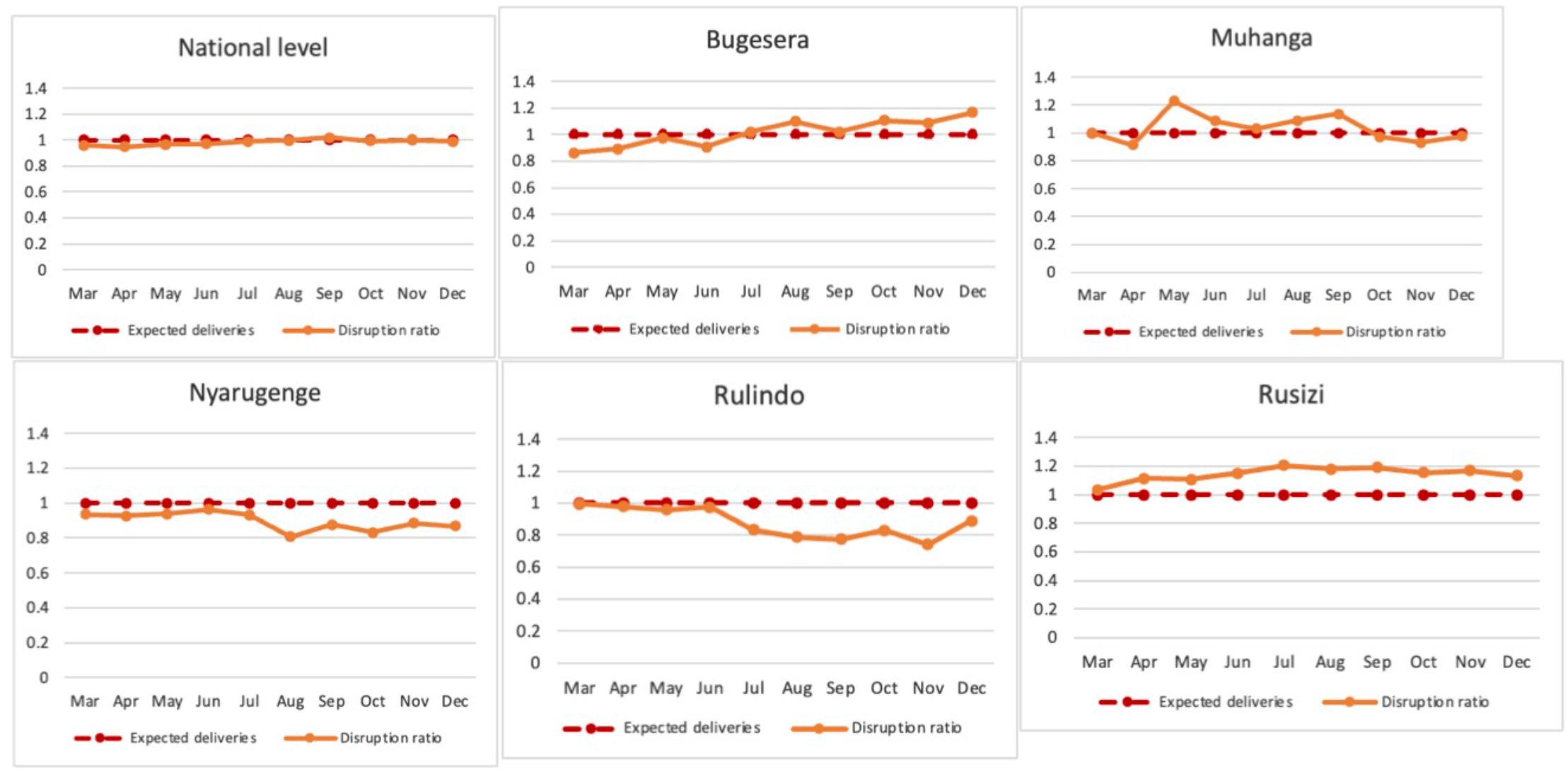
Facility-base delivery: Monthly disruption ratio for a selection of districts in Rwanda in 2020

Only five districts had lower numbers in individual months of facility-based deliveries than were expected. Nyamagabe and Nyarugenge had lower numbers of deliveries from March to December 2020 than expected. Rulindo had lower numbers of deliveries from July to December 2020 than expected. In Bugesera, there were lower numbers of facility-based deliveries than expected between March and July 2020, and greater numbers than expected between September and December 2020.

Based on key informant interviews, it is likely that implementation strategies such as building on the community health worker program and community-based health care delivery, establishment of maternal waiting areas in some areas, and provision of transport for both patients and health care providers, had a positive impact in terms of maintaining access to and demand for facility-based delivery services.

### Four or more antenatal care visits (ANC4+)

Cumulative disruption: Bugesera, Gasabo, Kicukiro, Ngoma, Ngororero, and Nyagatare had higher than expected disruptions to ANC4 services compared to other districts (Figure 5), with lower ANC4+ attendance than expected (disruption ratio <1). There was higher than expected ANC4+ attendance in 20 of Rwanda’s 30 districts (disruption ratio >1). In Burera, Gisagara, Muhanga, Nyabihu, and Rusizi, the disruption ratio was greater than 1.5, meaning substantially higher than expected ANC4+ attendance.

**Figure 5.**
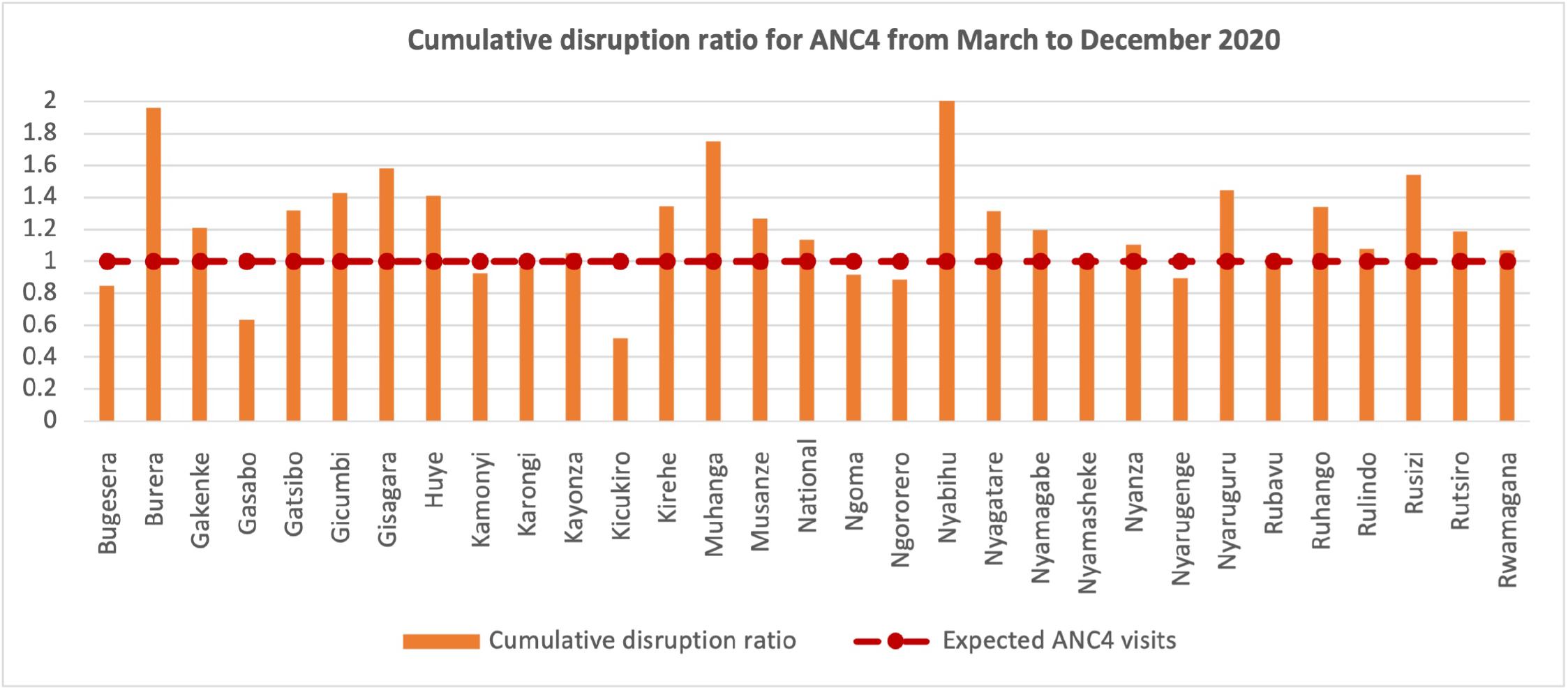
Monthly cumulative disruption ratios of four or more antenatal care visits in Rwanda between March-December 2020

Monthly disruption: For monthly disruption (Figure 6), there were higher than expected ANC4+ visits in Burera, Nyamagabe, Nyagatare, Nyaruguru, Nyabihu, Muhanga, Kirehe, Huye, Gicumbi, Gisagara, Gatsibo, Ruhango, and Rusizi in 2020. A greater disruption for ANC4+ (<1) was seen in Kigali especially in Kicukiro and Gasabo districts compared to other districts. Figure 6 shows monthly disruption nationally and for one district in each of Rwanda’s five provinces for illustration; full district-level results are available in the supplemental materials.

**Figure 6.**
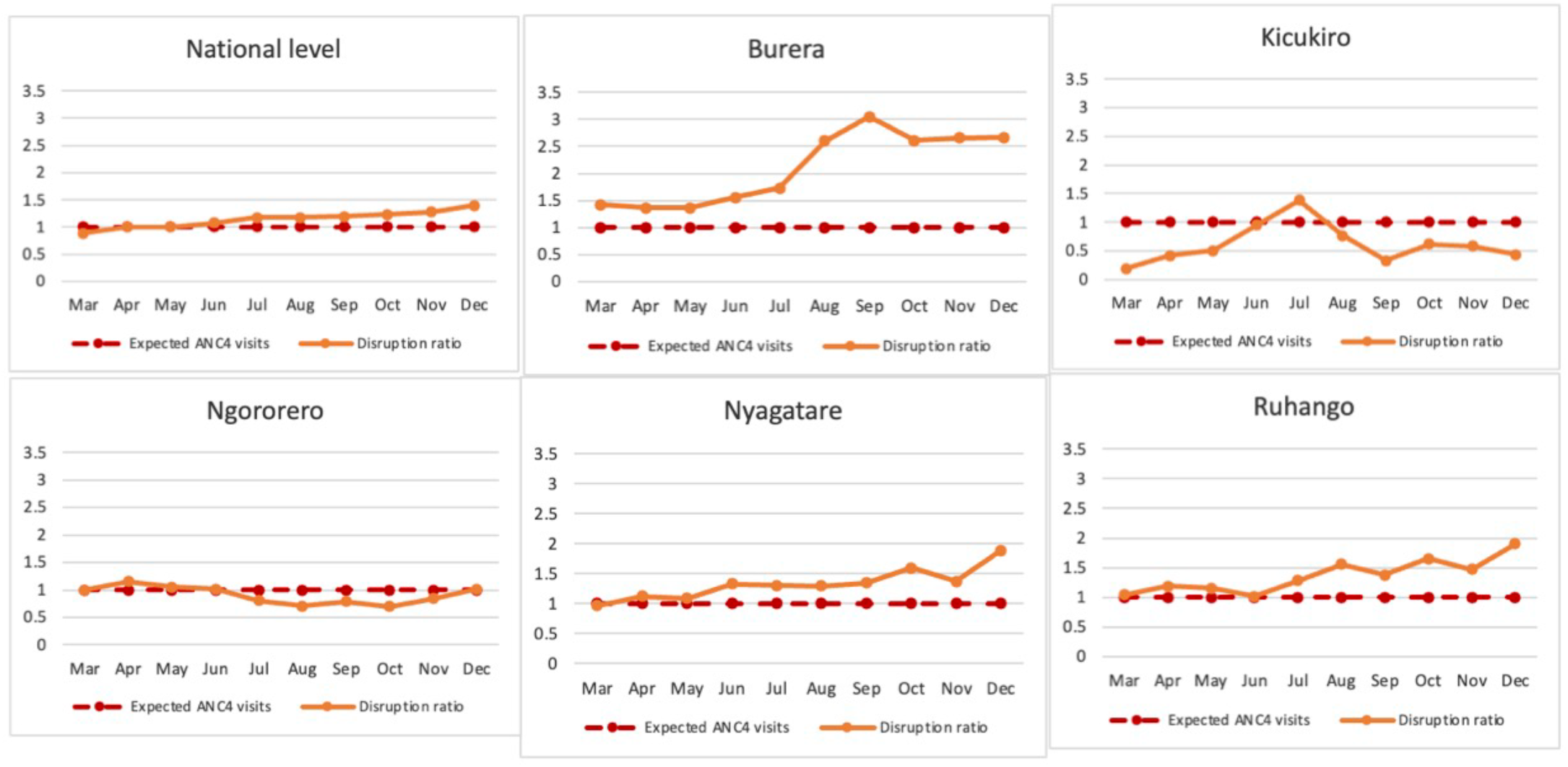
Four or more antenatal care visits: Monthly disruption ratio for a selection of districts in Rwanda in 2020

Based on key informant interviews, implementation strategies such as engagement and education of the community and community health worker interventions likely had a positive impact in terms of maintaining access to and demand for antenatal care services.

### Reported diarrheal cases

Cumulative disruption: There were cumulative disruptions to reported diarrheal cases in most districts from March to December 2020 (Figure 7). In nearly all districts the disruption ratio was <1, meaning that there were fewer than expected reported diarrhea cases. In Karongi, Kirehe, Nyabihu, Rubavu, Rusizi, and Rutsiro, the cumulative disruption ratio was >1, with greater than expected reported diarrheal cases between March to December 2020.

**Figure 7.**
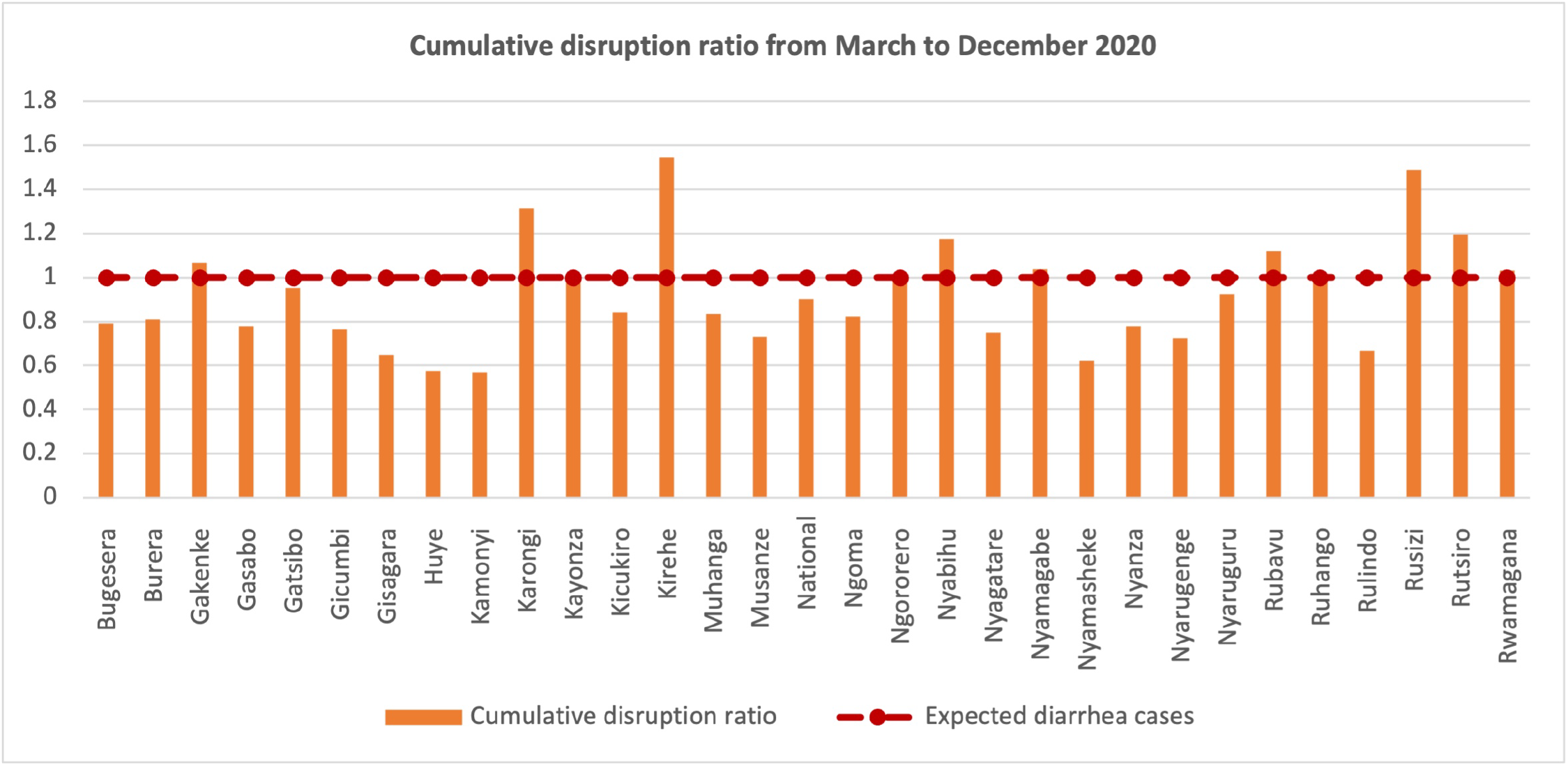
Monthly cumulative disruption ratios of reported diarrheal cases in Rwanda between March-December 2020

Monthly disruption: For monthly disruption (Figure 8), a number of districts had lower reported diarrheal cases than expected between March and July 2020 including Burera, Gakenke, Huye, Muhanga, and Ngorero. For districts with higher reported diarrheal cases than expected, this was often in the second half of the year (September to December 2020); this was the case in Karongi, Kirehe, Rusizi, Rutsiro, and Gakenke.

**Figure 8.**
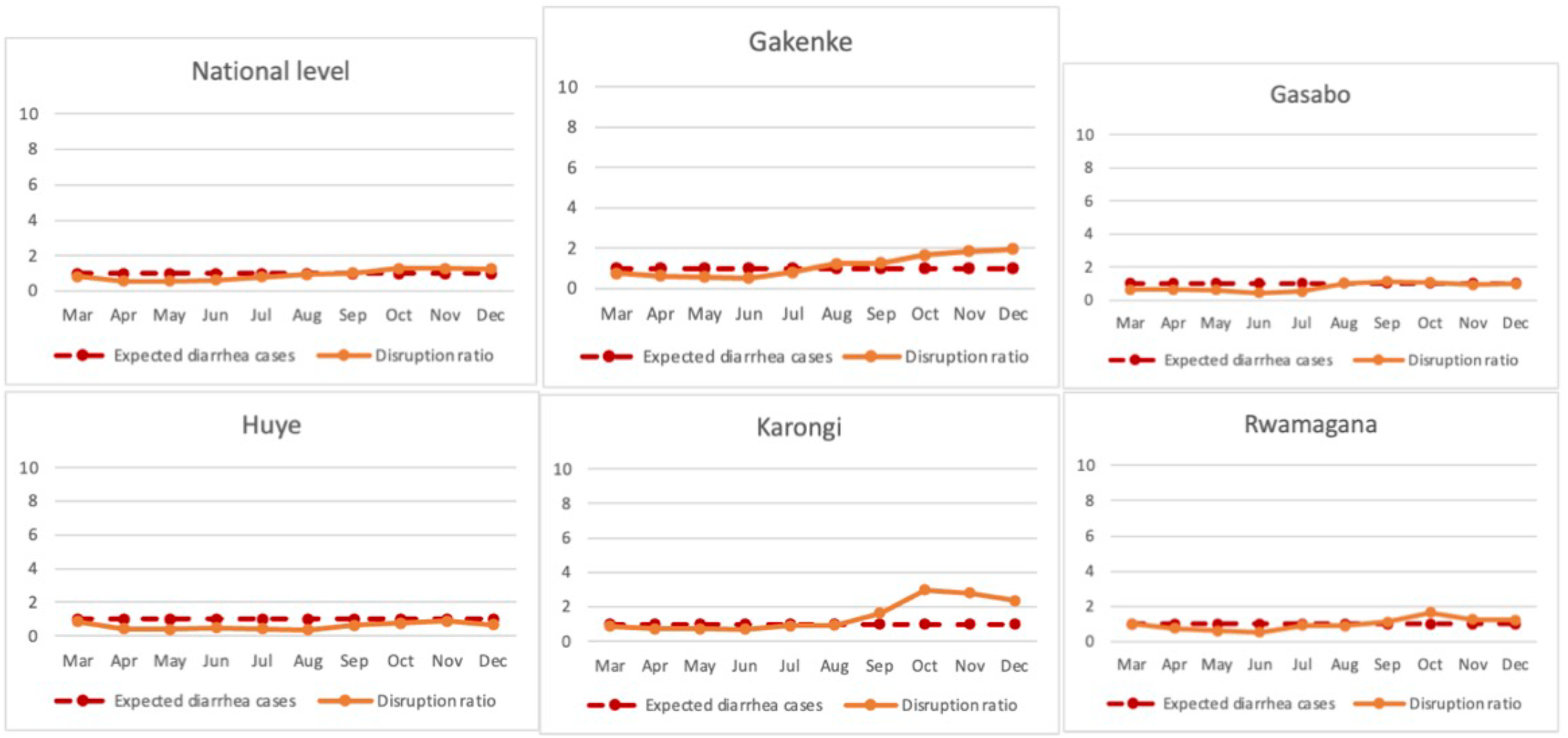
Reported diarrheal cases: Monthly disruption ratio for a selection of districts in Rwanda in 2020

Many of the districts experienced minimal disruption throughout March to December 2020, including Gasabo, Nyaruguru, Nyagatare, and Rwamagama. Figure 8 shows monthly disruption nationally and for one district in each of Rwanda’s five provinces for illustration; full district-level results are available in the supplemental materials.

Based on key informant interviews, it is likely that implementation strategies such as community health worker interventions, increases in water, sanitation, and hygiene (WASH) interventions to promote hygiene practices such as handwashing, as well as anecdotal reports of decreasing circulation of infectious diseases resulting from school closure and social distancing likely had in impact in reducing the numbers of diarrheal cases experienced and reported.

### Contextual factors supporting implementation of the evidence-based interventions

Based on key informant interviews, there was evidence of disruption nationally and subnationally during the earliest period of COVID-19. However, a majority of the key informants we spoke to agreed that the delivery of essential health services returned to normal levels soon after the national lockdown was lifted (on April 30, 2020). Key informants reasoned that the return to normal health service levels was due to the commitment to service delivery at all levels, strong community sensitization and instructions to health care workers and community health workers, and collaboration across partners to maintain or increase coverage. According to one key informant, *“Since we had to organize ourselves and make sure that services are delivered to our people, everything went on, despite the fact that we were in COVID*.*”*

Facilitators of implementation strategies targeting maintenance of facility-based delivery services and access and ANC4+ access and participation likely include Rwanda’s focus on equity, strong existing community health system including community health workers, preexisting culture of accountability, existing culture of learning and improvement, and resilient health system due to previous epidemic preparedness. According to key informants, maternal and child health community health workers would identify pregnant women and work with local leaders to track their status and sensitize the women to go to health centers for delivery (facilitator: strong existing community health system including community health workers). The importance of provision of transport during lockdown, both for providers and for patients, was one outcome of a focus on equity and existing system of accountability. For example, one key informant explained,

> “*For a people or a person who are in very remote area, normally here in Rwanda, local leaders have a means of transport. If there is no ambulance in the area, local leaders in this area helped us to bring the person who need medical care*…*we also increased our mobility of ambulances, even in the community, not only at health centers*.”

The existing culture of learning and improvement was important in the health system’s ability to adapt its strategies to respond to the disruptions during the first few months of the lockdown. One key informant noted that, “*In one month, we realized that the decrease in health seeking behavior, we designed a strategy for the community to seek treatment to CHWs [community health workers]*.”

Rwanda established both national and district level Public Health Emergency Operations Centers at the beginning of the pandemic, in addition to provincial level command posts, in July 2020, to further decentralize the country’s outbreak response measures.[15] The National Joint Task Force, composed of multidisciplinary teams and supported by a subnational task force from all 30 districts, was established to ensure the coordination around COVID-19 and support health care delivery. Command posts and the use of district-level versus national level lockdowns for pandemic control measures (facilitator: resilient health system due to previous epidemic preparedness) helped to minimize disruption and maintain access to and delivery of these essential health services across districts. One key informant explained,

> *“The command post played the role in the coordination of the logistics, ensuring the availability of cars, transport for the people to get services especially people moving from one place to another…It played a big role in logistics and in information-sharing across the country to ensure people were really getting services*.*”*

In terms of reported diarrheal cases and maintaining access to diarrheal care, some of the factors related to COVID-19-specific response measures, including lockdown and movement restriction, and school closures, likely also positively impacted infectious disease patterns including decreases in the burden of diarrheal cases.

## Discussion

We found that subnational variability of disruptions of primary health care services was relatively minimal in Rwanda, and that in general districts across Rwanda were able to continue to provide essential health services with low levels of disruption during the first phase (March to December 2020) of COVID-19. We also found that implementation strategies such as command posts, community health worker interventions, community engagement and education, and provision of transport were important factors in delivering minimal disruption across most districts in Rwanda during the first phase of COVID-19. Rwanda’s focus on equity likely helped to strengthen facilitating contextual factors including a culture of accountability and a strong pre-existing community health system and structure, which contributed to the low level of disruption and minimal subnational variability in the interventions we considered.

The low disruption to, and in some cases actual increase in, ANC4+ visits and facility-based deliveries demonstrate the importance of community health workers for maintaining essential health services and for equity in service delivery of interventions. A recent maternal health study expounded on the important role the health system plays in maternal health.[12] In the experience of Rwanda, community health workers were a powerful avenue to build community trust, educate women, and encourage participation in interventions targeting reduced U5M. Additionally, the inclusion of community health workers as part of the pandemic response workforce, as well as a community-based prevention orientation of the health system, were found to be enablers of primary health care implementation in a scoping review of primary health care barriers and facilitators during COVID-19.[13]

The use of command posts was an indication of governance, leadership, and coordination; many countries in sub-Saharan Africa established national committees but a common experience was a lack of established Emergency Operation Centers[14]; this was a strength for Rwanda, which established national and district level Public Health Emergency Operations Centers at the beginning of the pandemic, and added provincial level command posts a few months later to further decentralize the country’s outbreak response measures.[15]

Rwanda’s focus on equity as a priority has been well documented [16–18], and the minimal subnational variability and subnational disruption suggests that a national, ongoing prioritization of equity in health care access and services might be an important contributor to these outcomes. Edelman and colleagues found that an equity focus was an important enabler of primary health care access and performance, and that use of a lens of equity was a key enabler of primary health care implementation and governance.[13] Some amount of the disruption ratio showed “positive” disruption, e.g. *more* coverage than expected – higher levels of facility-based deliveries, more women attending four or more antenatal care visits. This is unusual compared to findings in many other countries and regions during the same COVID-19 period; decreases in facility-based deliveries and antenatal care visits were commonly noted.[19–24] This experience wasn’t entirely unprecedented, however, with a similar finding of increased facility-based deliveries during a similar period in Mozambique, though there was a longer lag from the onset of the pandemic to the increase in service provision.[25]

Rwanda’s low subnational variability for the disruption ratio, and low levels of disruption in general across districts, is reflective of the country’s resilient health system. We believe that the capacity to conduct both national and subnational analysis of disruption is valuable to generating meaningful lessons learned for communities in other regions and countries. Much of the provision of health care services, and, increasingly, decision-making, happens at the community level, increasing the value of comparative, subnational studies to help identify and localize critical approaches adopted in one community that could be adopted in others. Future research would additionally benefit from in-depth interviews with more local providers and representatives for deeper insights on decisions or interventions specific to individual communities and districts in Rwanda.

Our study had several limitations. The first is that community member representation, which would have provided valuable insight in our key informant interviews, was not possible due to time and resources constraints. The shorter duration of the period of COVID-19 included in this work limited our ability to detect transient changes in essential health service delivery and coverage. We focused only on the specific evidence-based interventions described in this article and the variability/disruption for other U5M-related evidence-based interventions might be different. The lower reported cases of diarrheal diseases during the period of COVID-19 in the study could be due to COVID-19 related measures and decreased treatment-seeking behavior. However, we were not able to investigate whether this reduction was from a reduction in incidence of diseases due to WASH-related hand washing practices and school closures leading to lower transmission of communicable diseases.

## Conclusion

We found minimal disruption across most districts in Rwanda, similar to previous findings of minimal disruption for the country as a whole, in the first phase of the COVID-19 pandemic and, furthermore, we found minimal subnational variability across districts. Implementation strategies such as community health worker interventions, community engagement and education, provision of transport, and command posts, were important factors in delivering minimal disruption across most districts in Rwanda during the first period of COVID-19. Rwanda’s focus on equity likely helped to strengthen facilitating contextual factors including a culture of accountability and a strong pre-existing community health system and structure, which contributed to the low level of disruption and minimal subnational variability in the interventions we considered. Rwanda’s experience offers potentially transferable knowledge for policymakers and decision-makers in other regions and countries working to decrease subnational inequities and minimize disruptions to essential health services during future health shocks.

## Author contributions

AA, LRH, and AB developed the concept. AA, AV, JTN, FS, ET, LRH, and AB designed and conducted the country case study research and contributed to the data analysis. AA, FS, AU, AV, JTN, ET, AK, LRH, and AB interpreted the results to form the manuscript. All authors contributed to writing and revision of the manuscript and had access to the data. All authors have approved the final version.

## Acknowledgements

We would like to thank the Institute for Health Metrics and Evaluation for methodological and data analysis input and guidance. We would also like to acknowledge and thank the key informants and stakeholders from partner organizations and national and subnational health institutions for providing valuable insights.

## Supplemental material

Supplemental figure 1. Monthly disruption ratio of facility-base delivery in Rwanda in 2020, by district

Supplemental figure 2. Monthly disruption ratio of four or more instances of antenatal care visits (ANC4+) in Rwanda in 2020, by district

Supplemental figure 3. Monthly disruption ratio of diarrheal cases reported at the community level in Rwanda in 2020, by district

## Data accessibility statement

The data that support the findings of this study are available from the Rwandan Ministry of Health (requests may be sent to). However, restrictions apply to the availability of these data, which were used for the current study, and so are not publicly available. The datasets used and/or analyzed during the current study are available from the corresponding author on reasonable request and with permission of the Rwandan Ministry of Health.

