## Supplemental materials for "Subnational equity in the delivery of primary health care interventions during health shocks: lessons learned from an implementation research study in Rwanda"

**Supplemental figure 1. Monthly disruption ratio of facility-base delivery in Rwanda in 2020, by district**

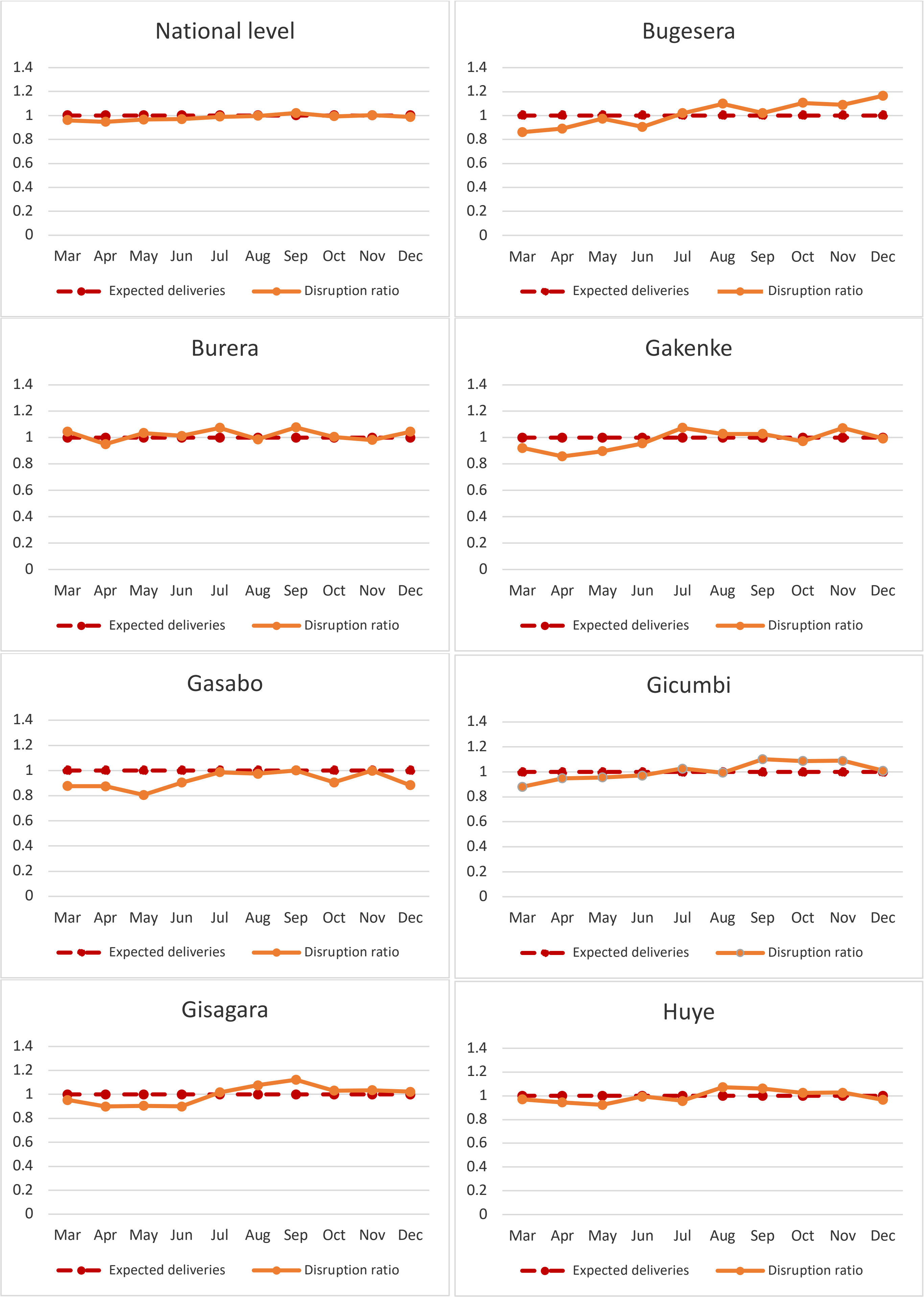

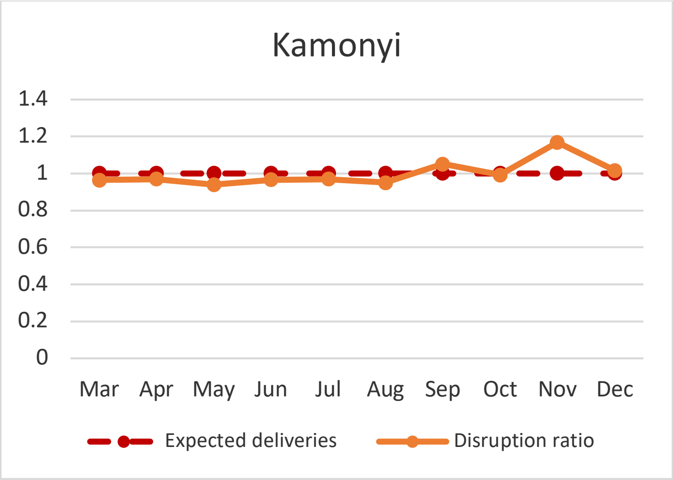
Kamonyi
1.4
1.2
1
0.8
0.6
0.4
0.2
0
Mar Apr May Jun Jul Aug Sep Oct Nov Dec
Expected deliveries
Disruption ratio

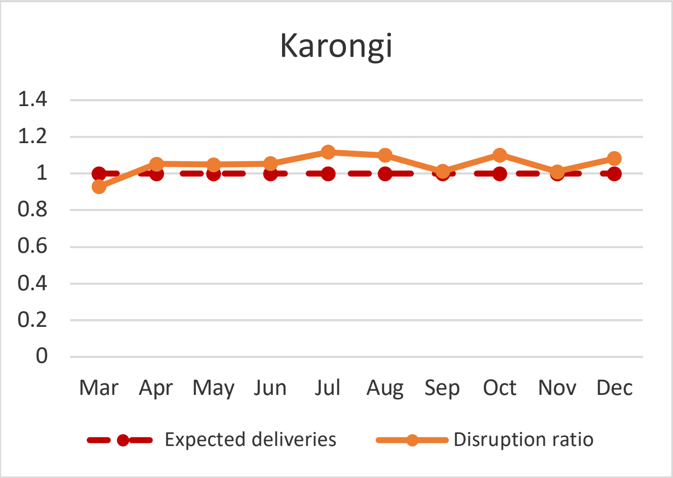
Karongi
1.4
1.2
1
0.8
0.6
0.4
0.2
0
Mar Apr May Jun Jul Aug Sep Oct Nov Dec
Expected deliveries
Disruption ratio

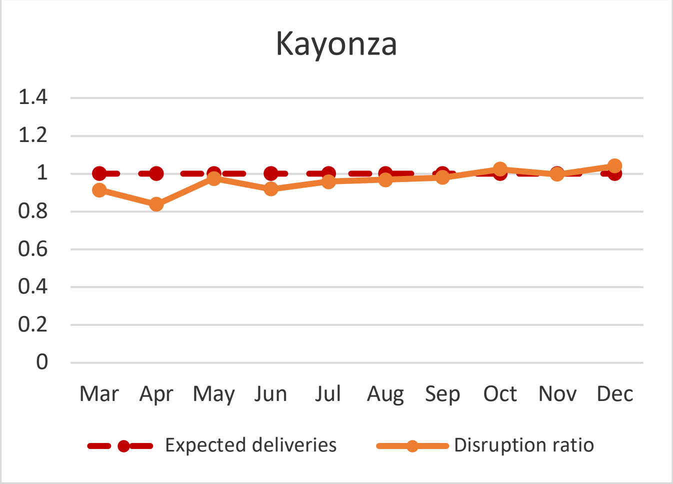
Kayonza
1.4
1.2
1
0.8
0.6
0.4
0.2
0
Mar Apr May Jun Jul Aug Sep Oct Nov Dec
Expected deliveries
Disruption ratio

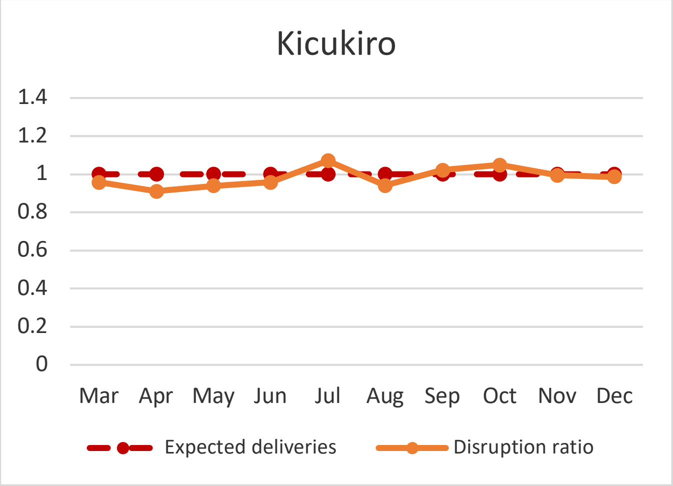
Kicukiro
1.4
1.2
1
0.8
0.6
0.4
0.2
0
Mar Apr May Jun Jul Aug Sep Oct Nov Dec
Expected deliveries
Disruption ratio

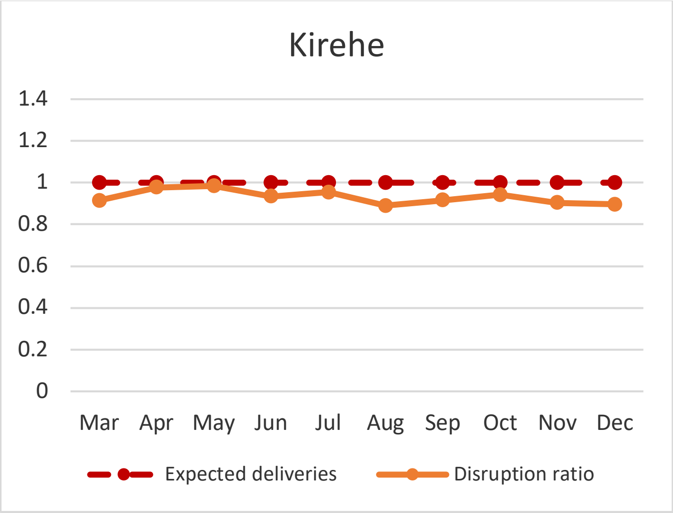
Kirehe
1.4
1.2
1
0.8
0.6
0.4
0.2
0
Mar Apr May Jun Jul Aug Sep Oct Nov Dec
Expected deliveries
Disruption ratio

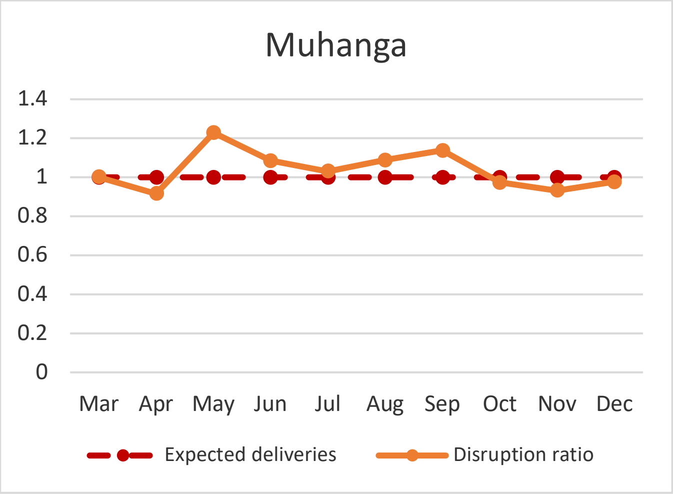
Muhanga
1.4
1.2
1
0.8
0.6
0.4
0.2
0
Mar Apr May Jun Jul Aug Sep Oct Nov Dec
Expected deliveries
Disruption ratio

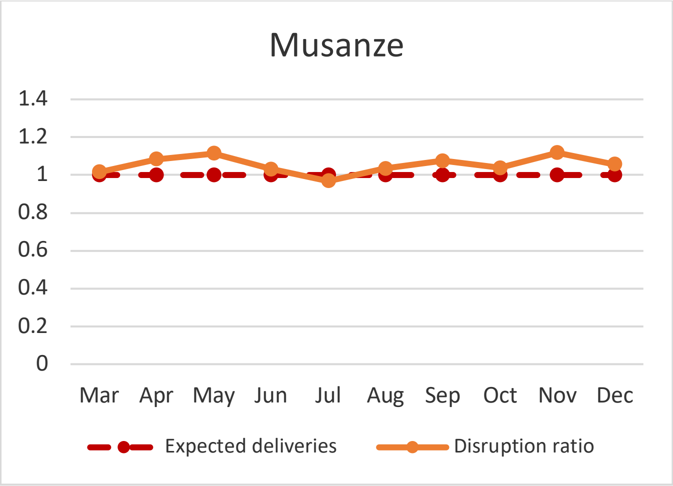
Musanze
1.4
1.2
1
0.8
0.6
0.4
0.2
0
Mar Apr May Jun Jul Aug Sep Oct Nov Dec
Expected deliveries
Disruption ratio

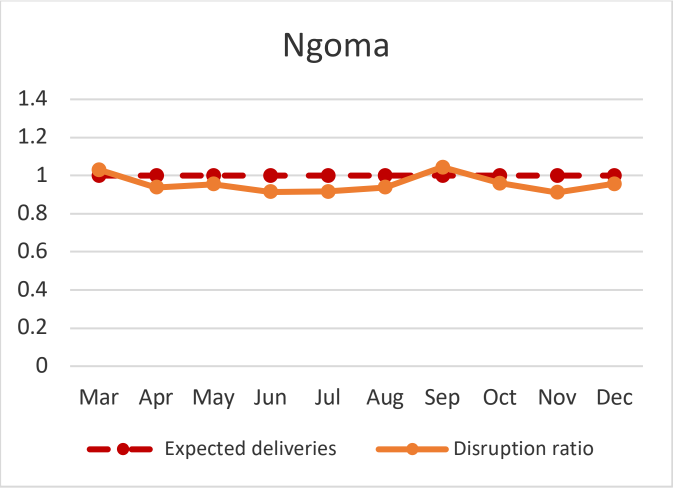
Ngoma
1.4
1.2
1
0.8
0.6
0.4
0.2
0
Mar Apr May Jun Jul Aug Sep Oct Nov Dec
Expected deliveries
Disruption ratio

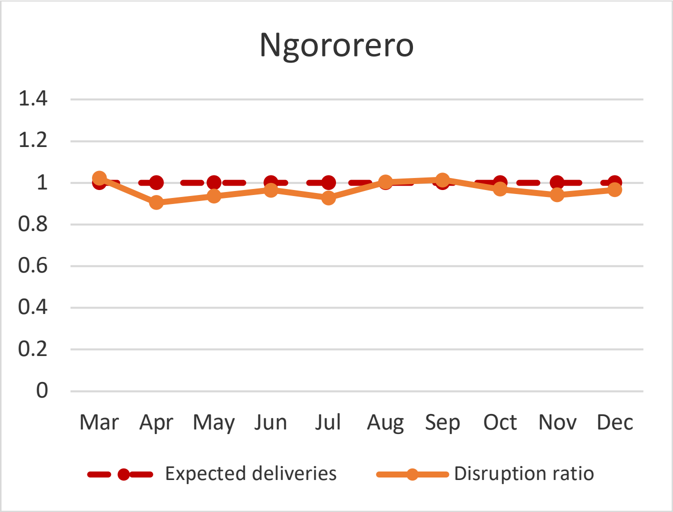
Ngororero
1.4
1.2
1
0.8
0.6
0.4
0.2
0
Mar Apr May Jun Jul Aug Sep Oct Nov Dec
Expected deliveries
Disruption ratio

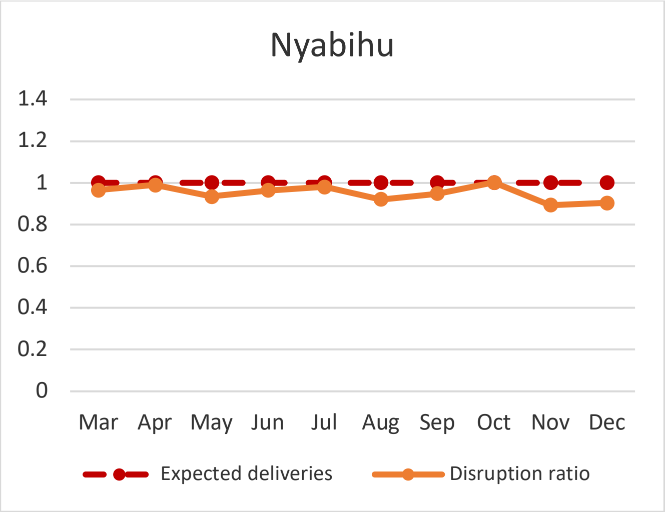
Nyabihu
1.4
1.2
1
0.8
0.6
0.4
0.2
0
Mar Apr May Jun Jul Aug Sep Oct Nov Dec
Expected deliveries
Disruption ratio

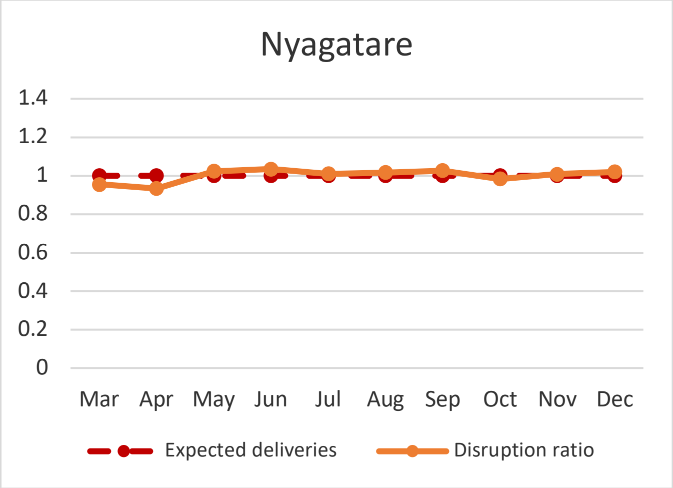
Nyagatare
1.4
1.2
1
0.8
0.6
0.4
0.2
0
Mar Apr May Jun Jul Aug Sep Oct Nov Dec
Expected deliveries
Disruption ratio

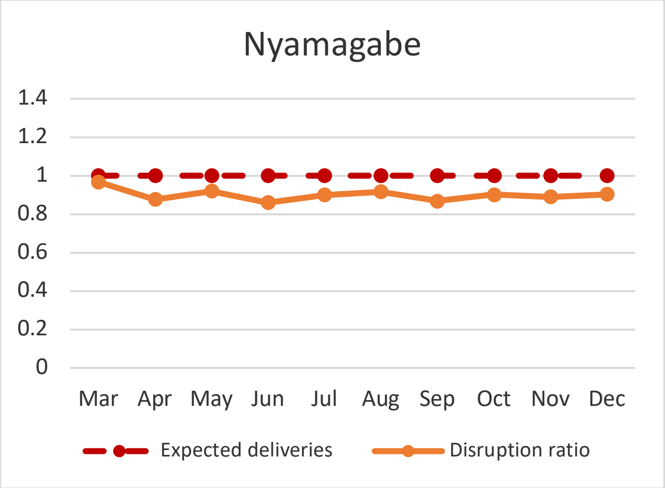
Nyamagabe
1.4
1.2
1
0.8
0.6
0.4
0.2
0
Mar Apr May Jun Jul Aug Sep Oct Nov Dec
Expected deliveries
Disruption ratio

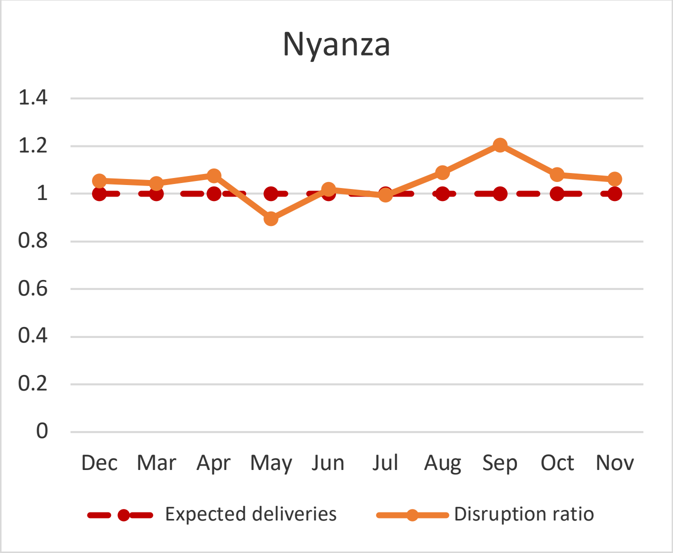
Nyanza
1.4
1.2
1
0.8
0.6
0.4
0.2
0
Dec Mar Apr May Jun Jul Aug Sep Oct Nov
Expected deliveries
Disruption ratio

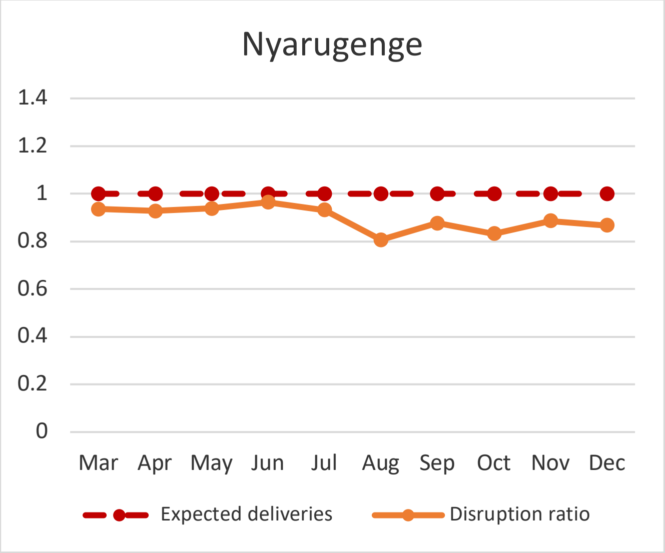
Nyarugenge
1.4
1.2
1
0.8
0.6
0.4
0.2
0
Mar Apr May Jun Jul Aug Sep Oct Nov Dec
Expected deliveries
Disruption ratio

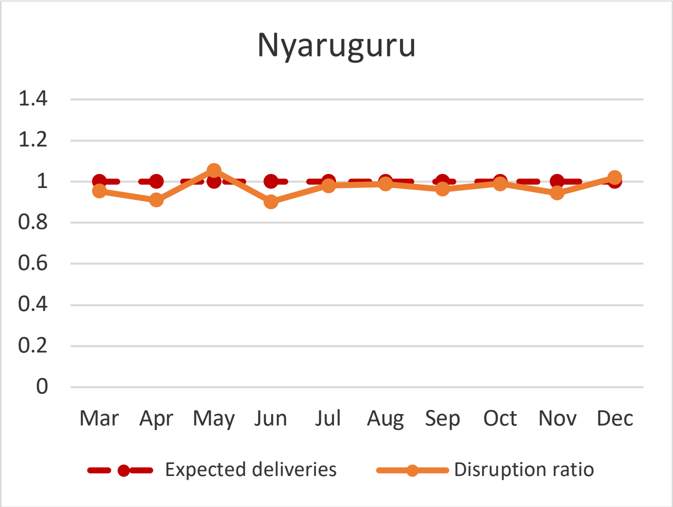
Nyaruguru
1.4
1.2
1
0.8
0.6
0.4
0.2
0
Mar Apr May Jun Jul Aug Sep Oct Nov Dec
Expected deliveries
Disruption ratio

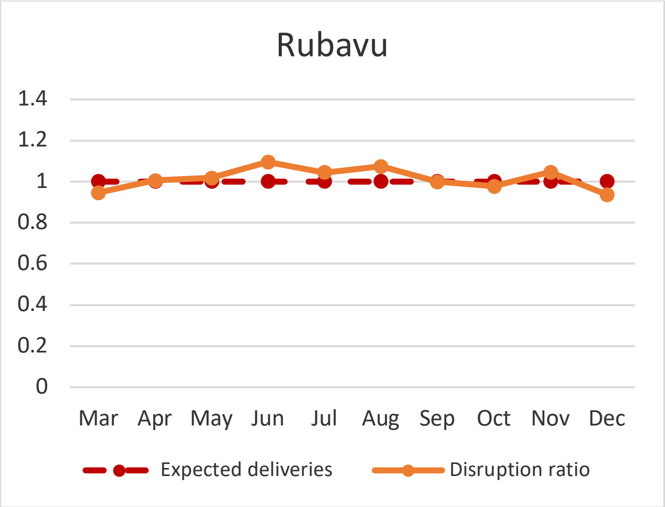
Rubavu
1.4
1.2
1
0.8
0.6
0.4
0.2
0
Mar Apr May Jun Jul Aug Sep Oct Nov Dec
Expected deliveries
Disruption ratio

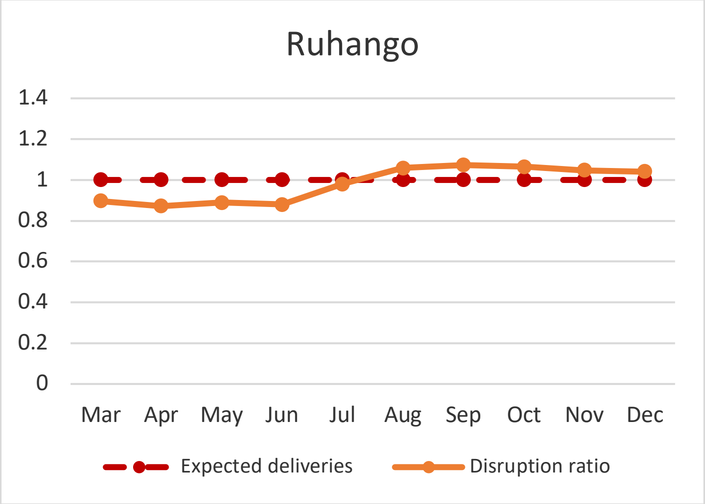
Ruhango
1.4
1.2
1
0.8
0.6
0.4
0.2
0
Mar Apr May Jun Jul Aug Sep Oct Nov Dec
Expected deliveries
Disruption ratio

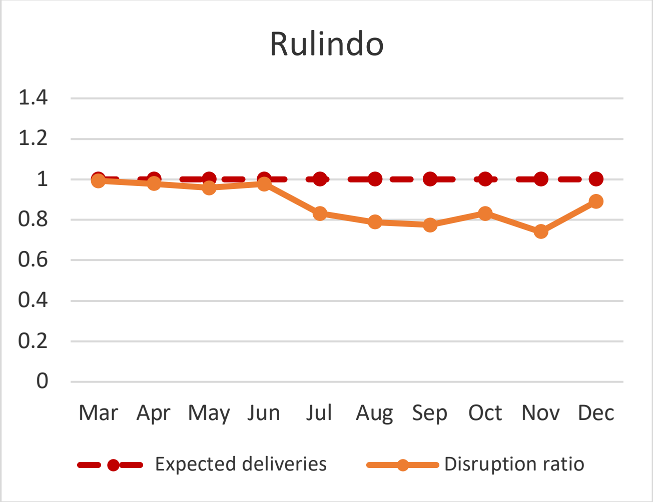
Rulindo
1.4
1.2
1
0.8
0.6
0.4
0.2
0
Mar Apr May Jun Jul Aug Sep Oct Nov Dec
Expected deliveries
Disruption ratio

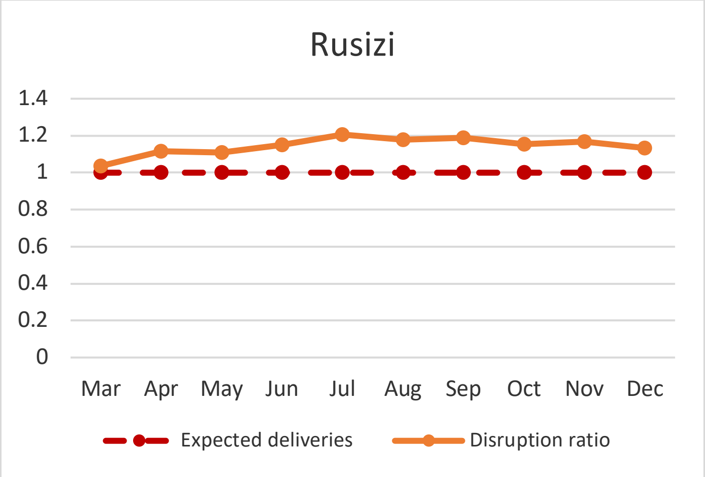
Rusizi
1.4
1.2
1
0.8
0.6
0.4
0.2
0
Mar Apr May Jun Jul Aug Sep Oct Nov Dec
Expected deliveries
Disruption ratio

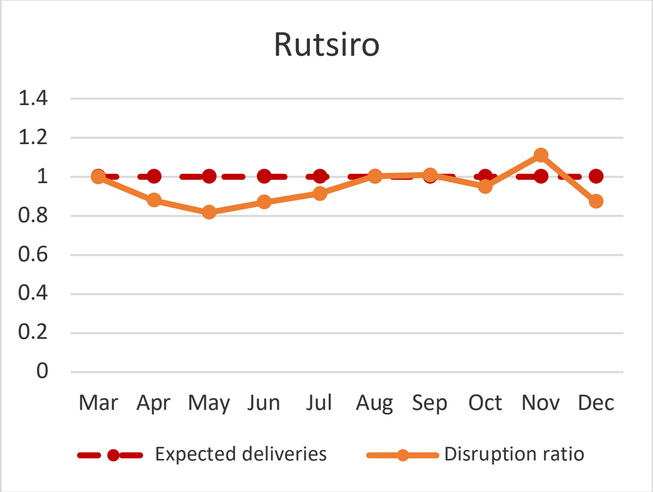
Rutsiro
1.4
1.2
1
0.8
0.6
0.4
0.2
0
Mar Apr May Jun Jul Aug Sep Oct Nov Dec
Expected deliveries
Disruption ratio

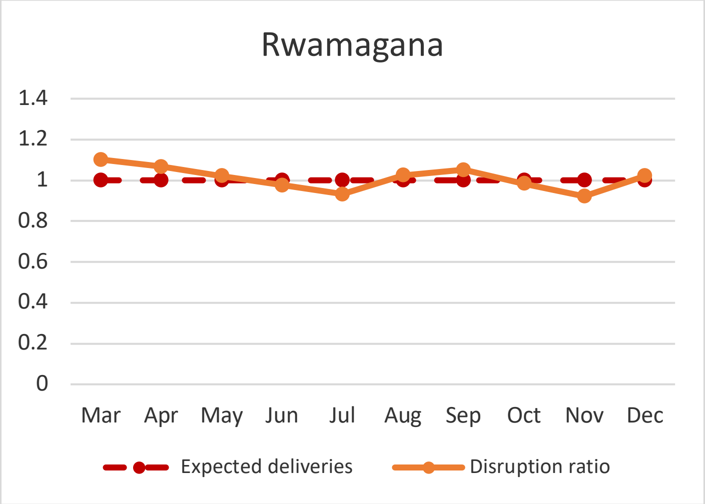
Rwamagana
1.4
1.2
1
0.8
0.6
0.4
0.2
0
Mar Apr May Jun Jul Aug Sep Oct Nov Dec
Expected deliveries
Disruption ratio

**Supplemental figure 2. Monthly disruption ratio of four or more instances of antenatal care visits (ANC4+) in Rwanda in 2020, by district**

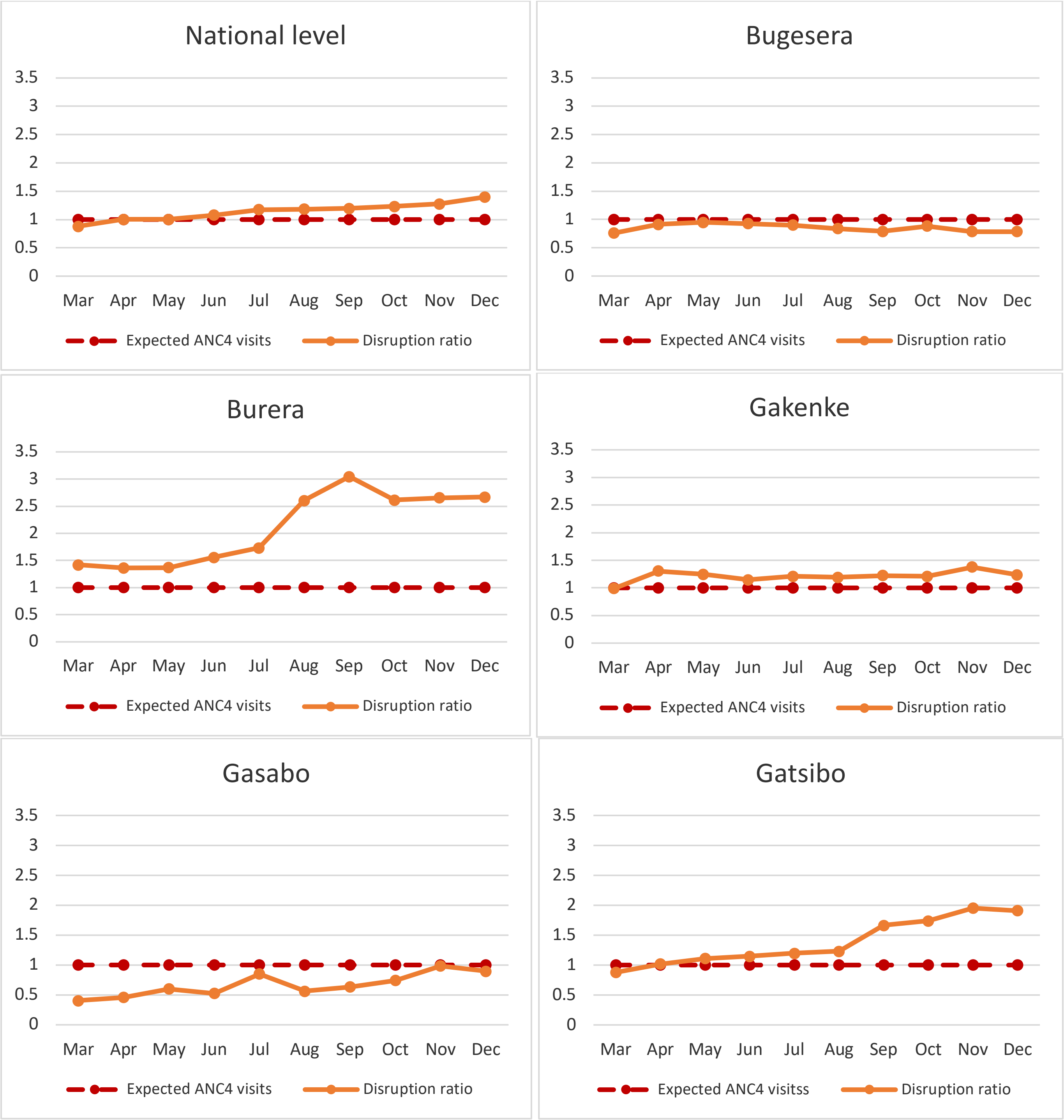

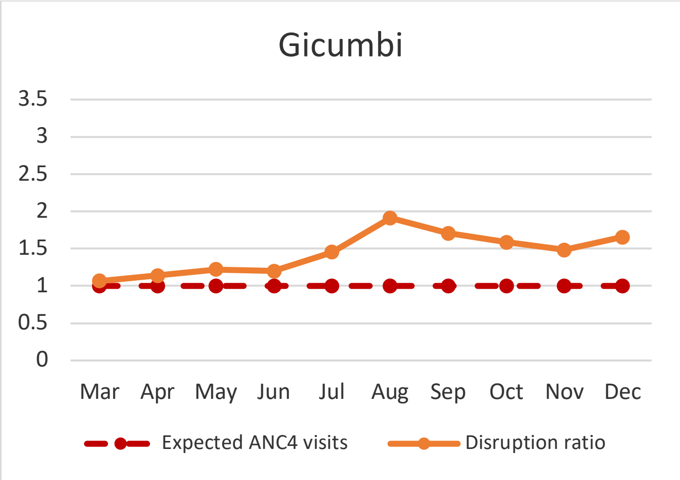
Gicumbi
3.5
3
2.5
2
1.5
1
0.5
0
Mar Apr May Jun Jul Aug Sep Oct Nov Dec
Expected ANC4 visits
Disruption ratio

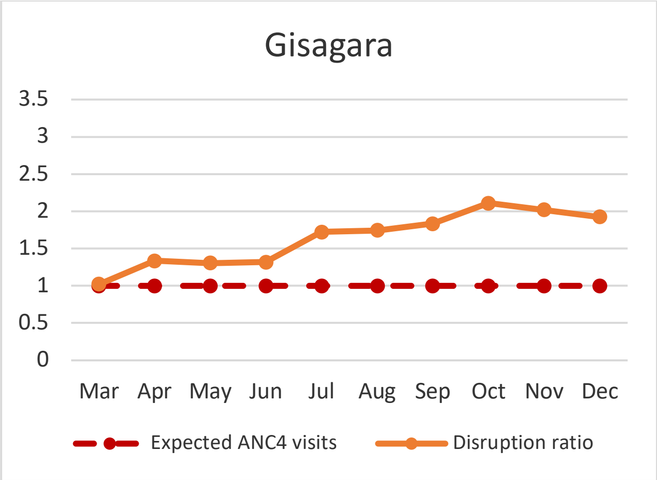
Gisagara
3.5
3
2.5
2
1.5
1
0.5
0
Mar Apr May Jun Jul Aug Sep Oct Nov Dec
Expected ANC4 visits
Disruption ratio

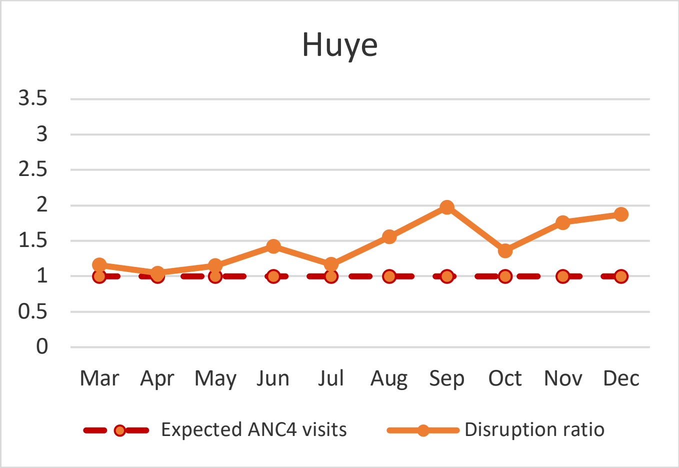
Huye
3.5
3
2.5
2
1.5
1
0.5
0
Mar Apr May Jun Jul Aug Sep Oct Nov Dec
Expected ANC4 visits
Disruption ratio

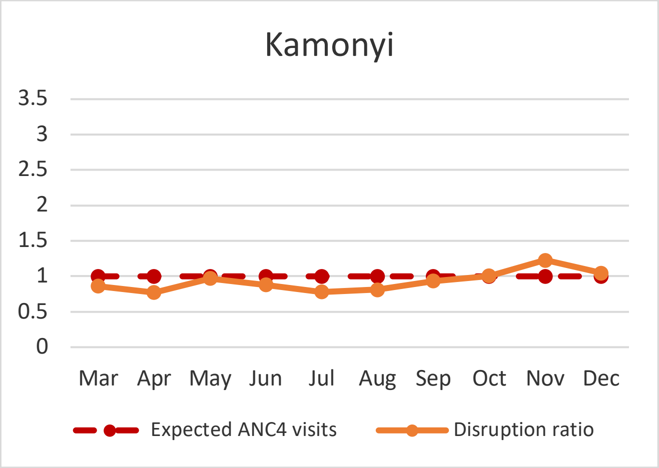
Kamonyi
3.5
3
2.5
2
1.5
1
0.5
0
Mar Apr May Jun Jul Aug Sep Oct Nov Dec
Expected ANC4 visits
Disruption ratio

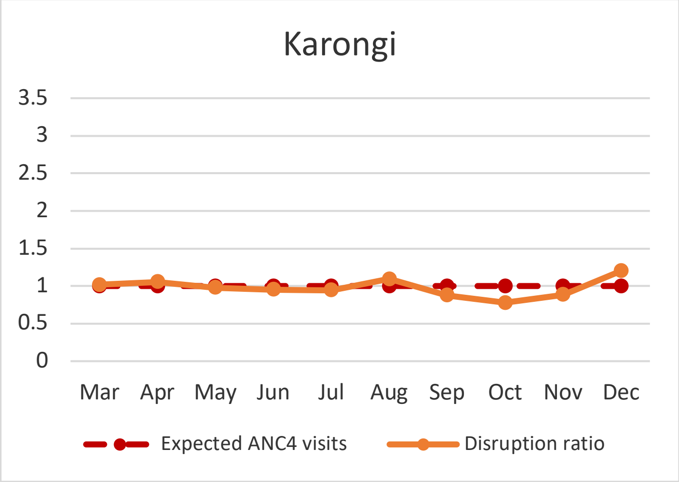
Karongi
3.5
3
2.5
2
1.5
1
0.5
0
Mar Apr May Jun Jul Aug Sep Oct Nov Dec
Expected ANC4 visits
Disruption ratio

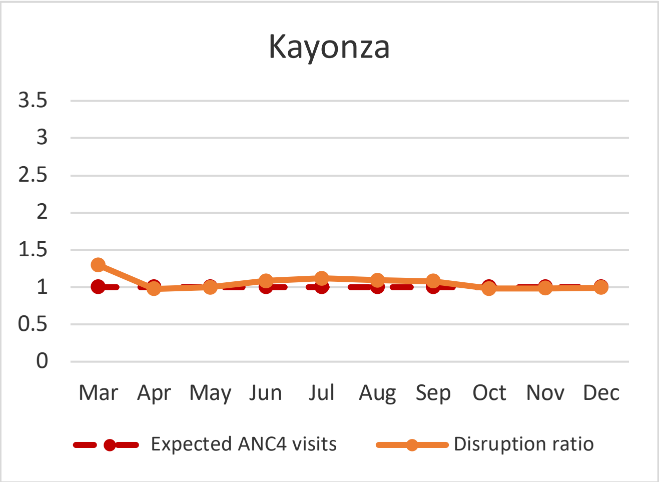
Kayonza
3.5
3
2.5
2
1.5
1
0.5
0
Mar Apr May Jun Jul Aug Sep Oct Nov Dec
Expected ANC4 visits
Disruption ratio

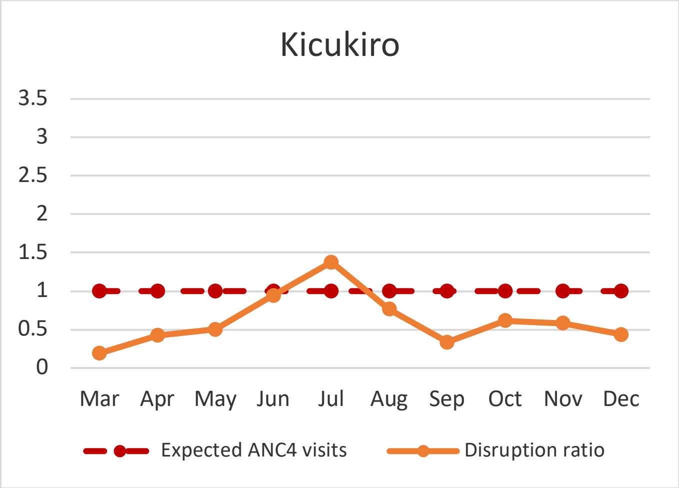
Kicukiro
3.5
3
2.5
2
1.5
1
0.5
0
Mar Apr May Jun Jul Aug Sep Oct Nov Dec
Expected ANC4 visits
Disruption ratio

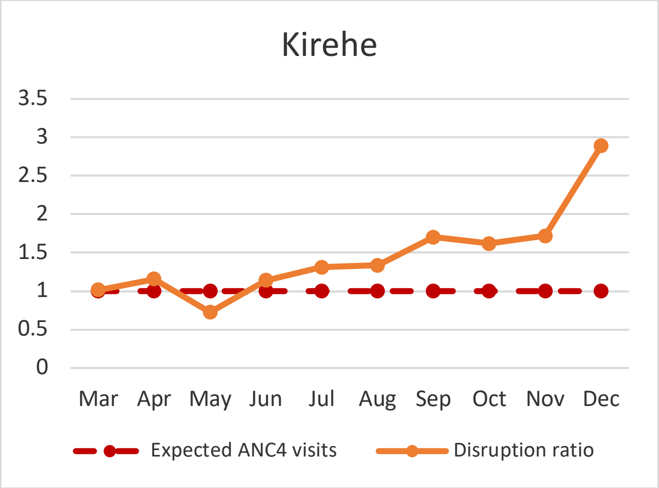
Kirehe
3.5
3
2.5
2
1.5
1
0.5
0
Mar Apr May Jun Jul Aug Sep Oct Nov Dec
Expected ANC4 visits
Disruption ratio

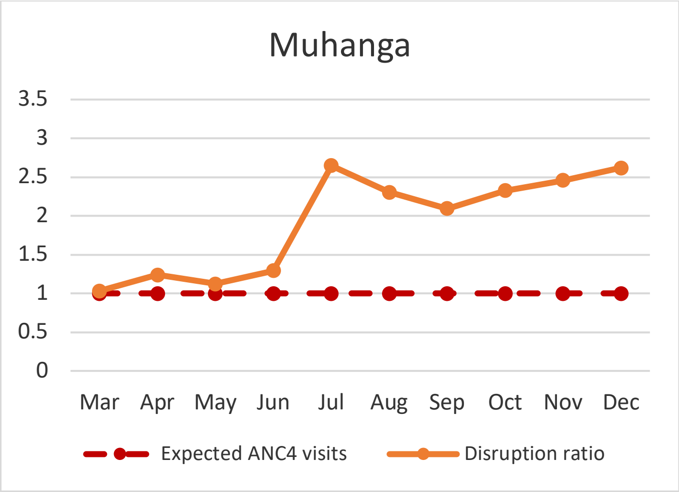
Muhanga
3.5
3
2.5
2
1.5
1
0.5
0
Mar Apr May Jun Jul Aug Sep Oct Nov Dec
Expected ANC4 visits
Disruption ratio

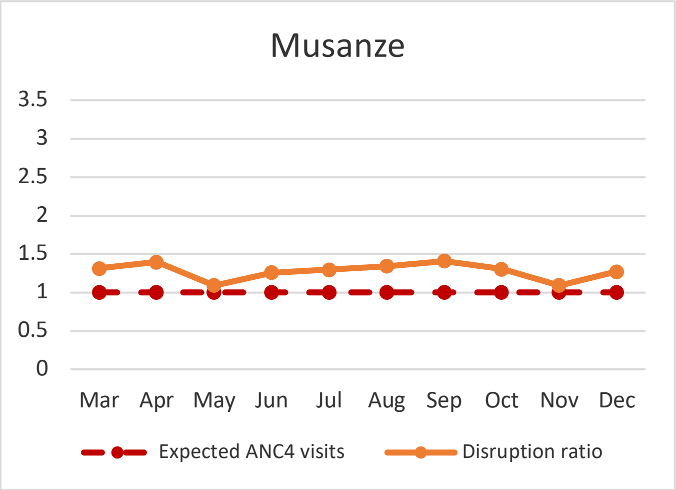
Musanze
3.5
3
2.5
2
1.5
1
0.5
0
Mar Apr May Jun Jul Aug Sep Oct Nov Dec
Expected ANC4 visits
Disruption ratio

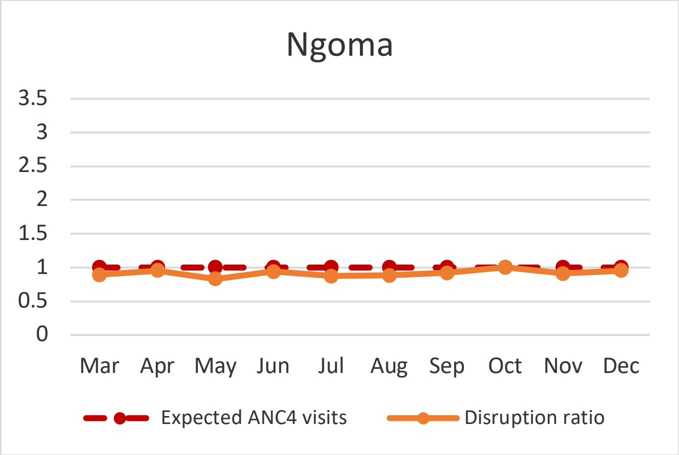
Ngoma
3.5
3
2.5
2
1.5
1
0.5
0
Mar Apr May Jun Jul Aug Sep Oct Nov Dec
Expected ANC4 visits
Disruption ratio

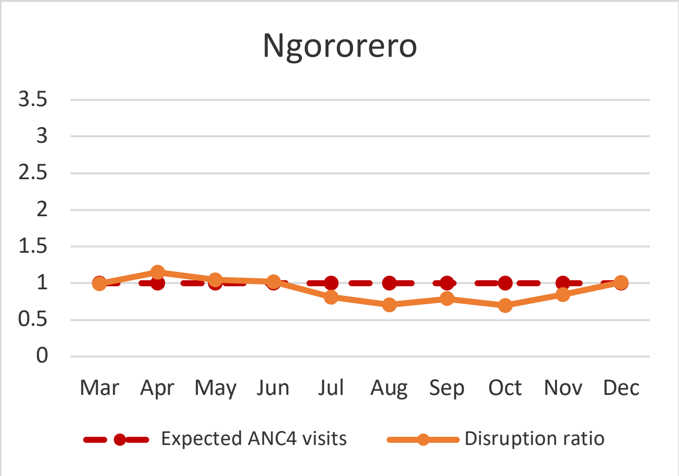
Ngororero
3.5
3
2.5
2
1.5
1
0.5
0
Mar Apr May Jun Jul Aug Sep Oct Nov Dec
Expected ANC4 visits
Disruption ratio

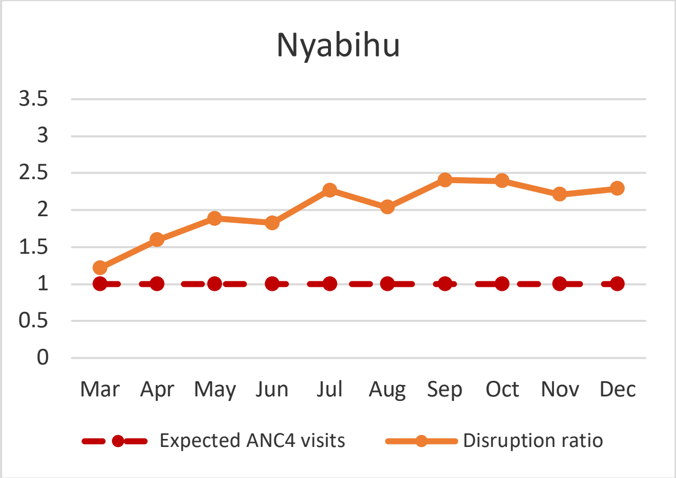
Nyabihu
3.5
3
2.5
2
1.5
1
0.5
0
Mar Apr May Jun Jul Aug Sep Oct Nov Dec
Expected ANC4 visits
Disruption ratio

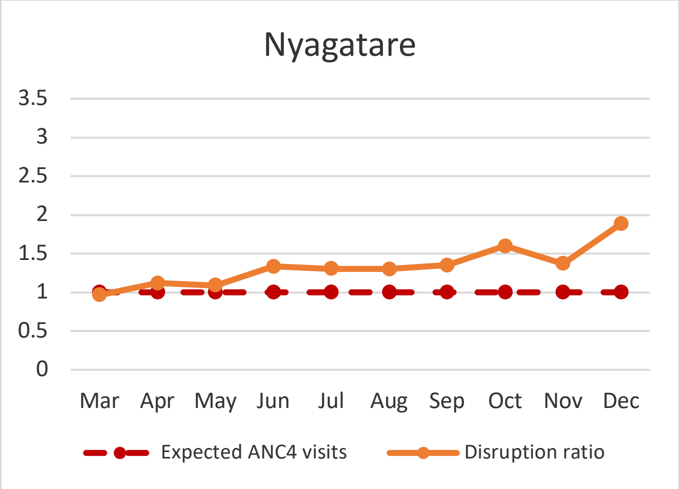
Nyagatare
3.5
3
2.5
2
1.5
1
0.5
0
Mar Apr May Jun Jul Aug Sep Oct Nov Dec
Expected ANC4 visits
Disruption ratio

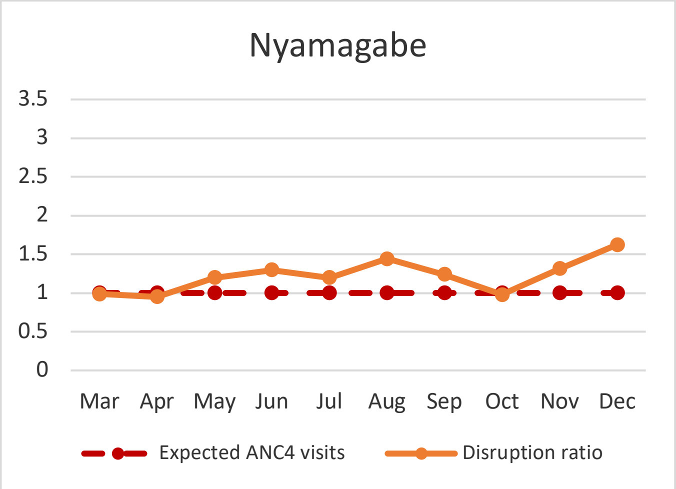
Nyamagabe
3.5
3
2.5
2
1.5
1
0.5
0
Mar Apr May Jun Jul Aug Sep Oct Nov Dec
Expected ANC4 visits
Disruption ratio

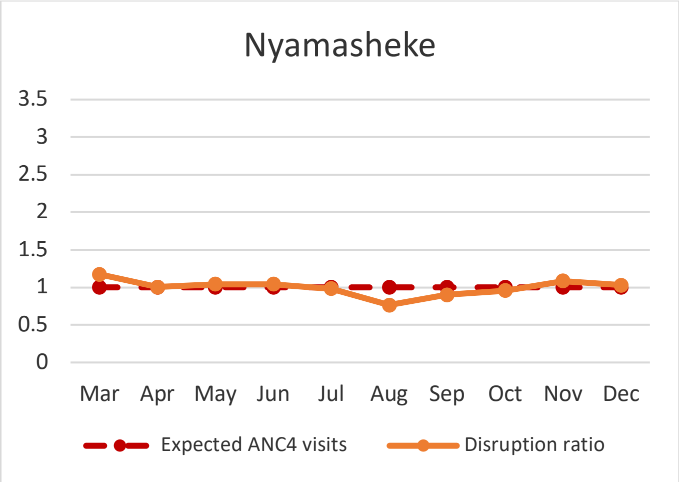
Nyamasheke
3.5
3
2.5
2
1.5
1
0.5
0
Mar Apr May Jun Jul Aug Sep Oct Nov Dec
Expected ANC4 visits
Disruption ratio

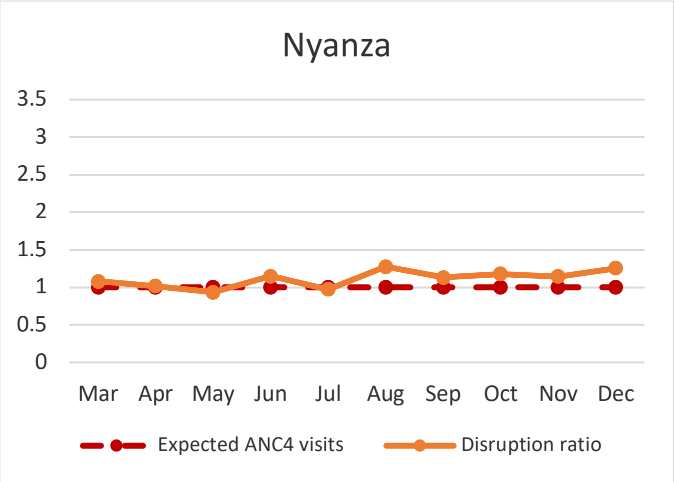
Nyanza
3.5
3
2.5
2
1.5
1
0.5
0
Mar Apr May Jun Jul Aug Sep Oct Nov Dec
Expected ANC4 visits
Disruption ratio

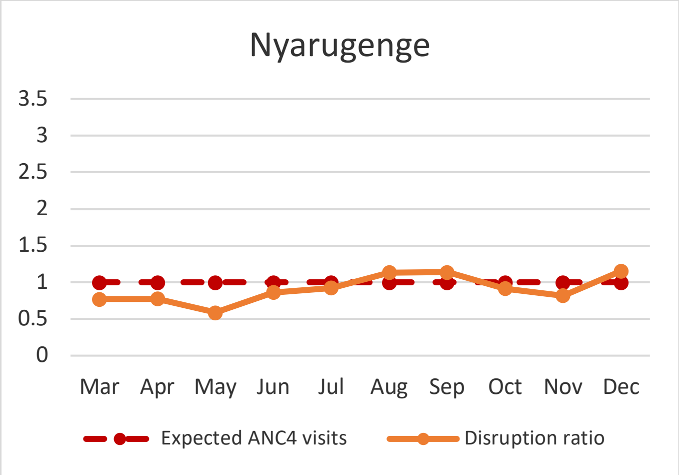
Nyarugenge
3.5
3
2.5
2
1.5
1
0.5
0
Mar Apr May Jun Jul Aug Sep Oct Nov Dec
Expected ANC4 visits
Disruption ratio

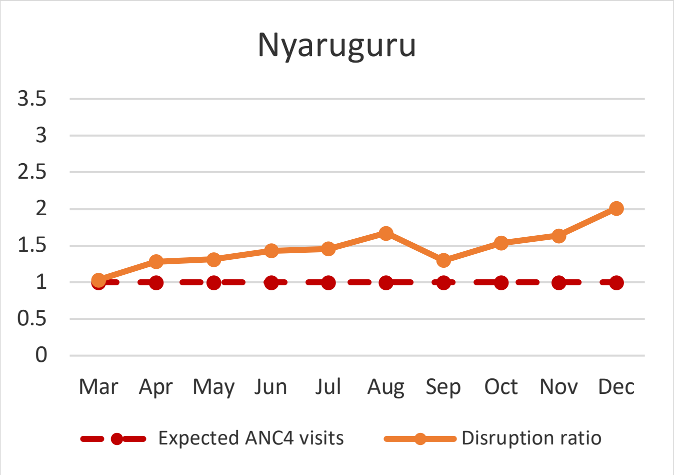
Nyaruguru
3.5
3
2.5
2
1.5
1
0.5
0
Mar Apr May Jun Jul Aug Sep Oct Nov Dec
Expected ANC4 visits
Disruption ratio

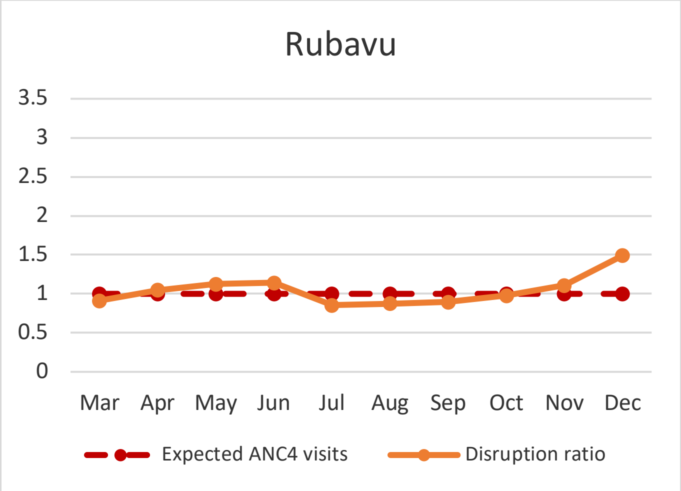
Rubavu
3.5
3
2.5
2
1.5
1
0.5
0
Mar Apr May Jun Jul Aug Sep Oct Nov Dec
Expected ANC4 visits
Disruption ratio

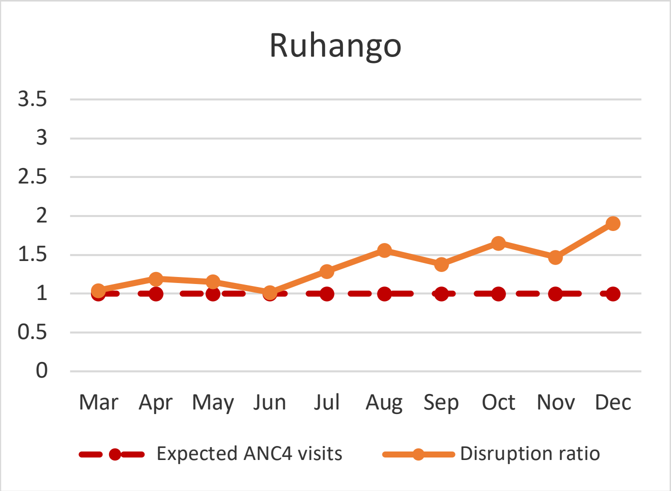
Ruhango
3.5
3
2.5
2
1.5
1
0.5
0
Mar Apr May Jun Jul Aug Sep Oct Nov Dec
Expected ANC4 visits
Disruption ratio

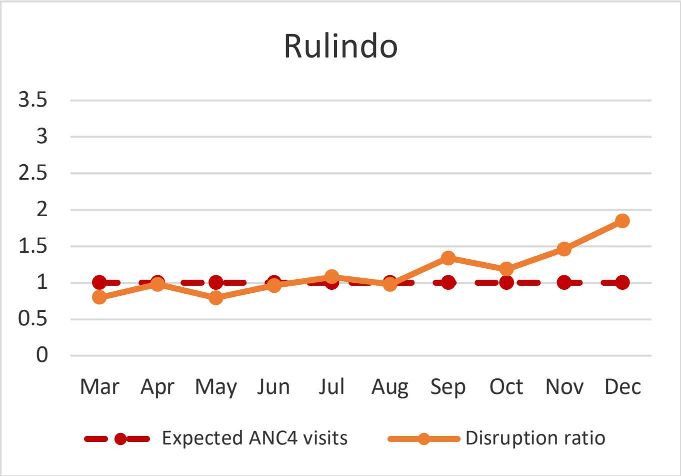
Rulindo
3.5
3
2.5
2
1.5
1
0.5
0
Mar Apr May Jun Jul Aug Sep Oct Nov Dec
Expected ANC4 visits
Disruption ratio

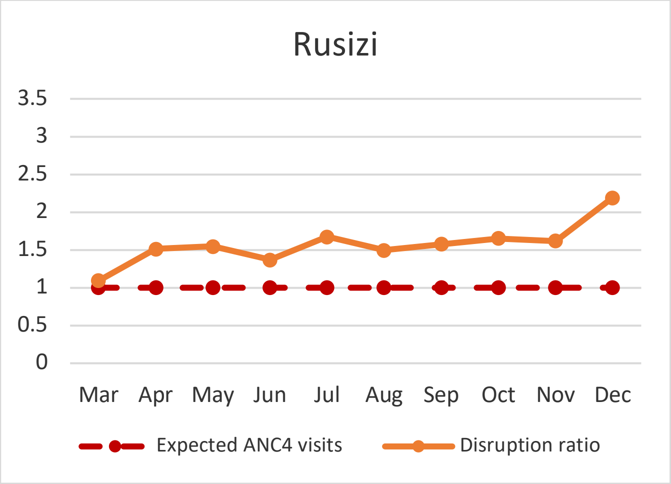
Rusizi
3.5
3
2.5
2
1.5
1
0.5
0
Mar Apr May Jun Jul Aug Sep Oct Nov Dec
Expected ANC4 visits
Disruption ratio

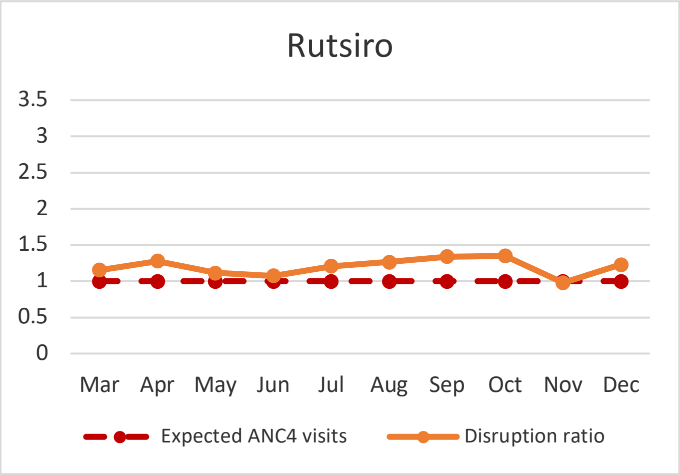
Rutsiro
3.5
3
2.5
2
1.5
1
0.5
0
Mar
Apr
May
Jun
Jul
Aug
Sep
Oct
Nov
Dec
Expected ANC4 visits
Disruption ratio

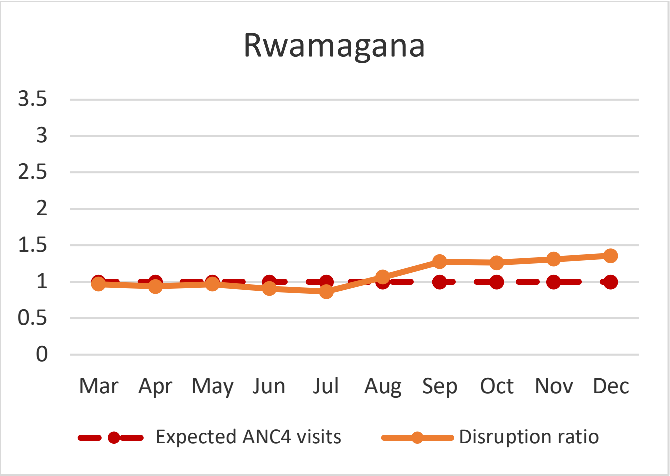
Rwamagana
3.5
3
2.5
2
1.5
1
0.5
0
Mar
Apr
May
Jun
Jul
Aug
Sep
Oct
Nov
Dec
Expected ANC4 visits
Disruption ratio

**Supplemental figure 3. Monthly disruption ratio of diarrheal cases reported at the community level in Rwanda in 2020, by district**

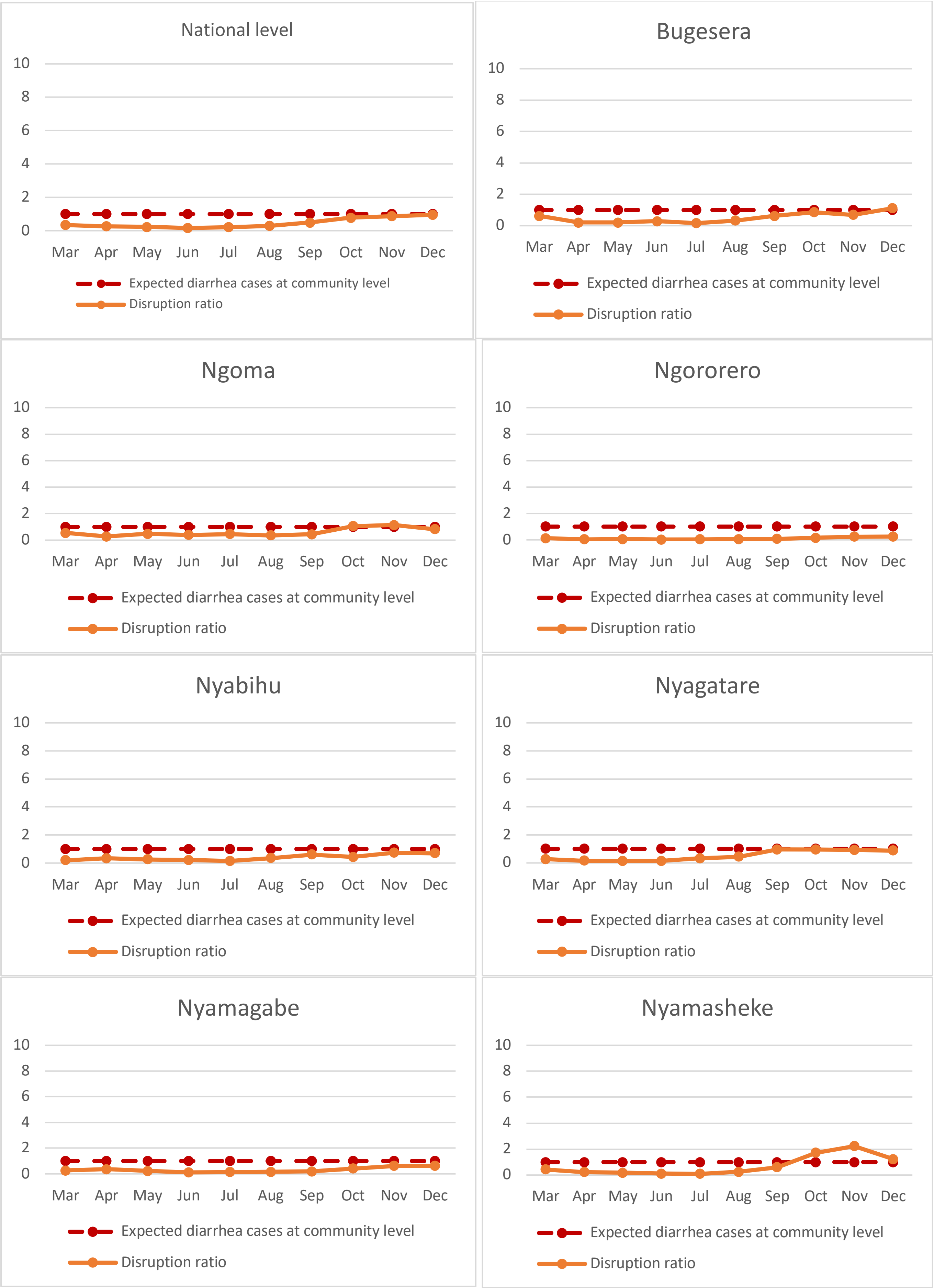

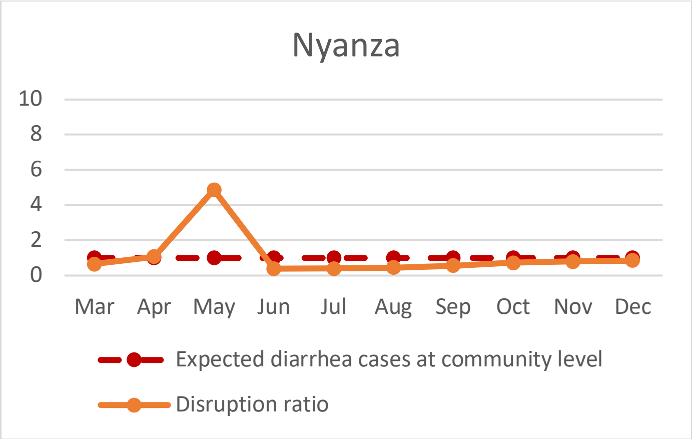
Nyanza
10
8
6
4
2
0
Mar Apr May Jun Jul Aug Sep Oct Nov Dec
Expected diarrhea cases at community level
Disruption ratio

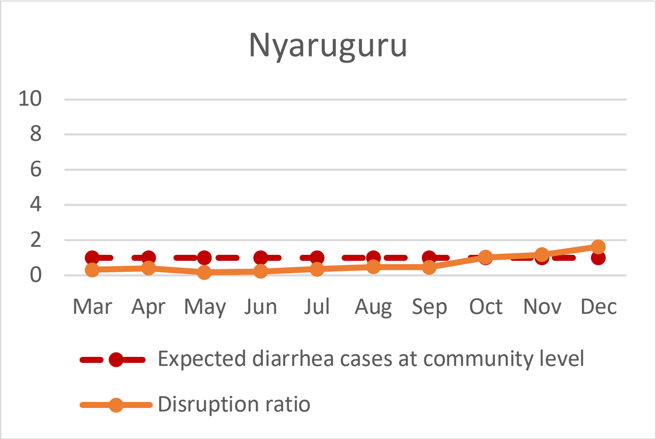
Nyaruguru
10
8
6
4
2
0
Mar Apr May Jun Jul Aug Sep Oct Nov Dec
Expected diarrhea cases at community level
Disruption ratio

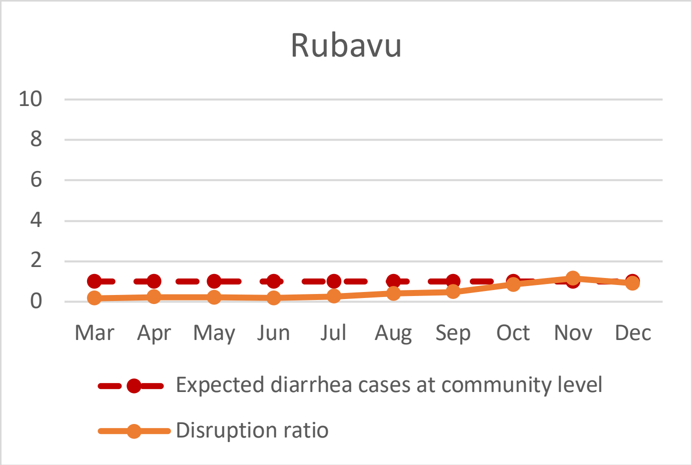
Rubavu
10
8
6
4
2
0
Mar Apr May Jun Jul Aug Sep Oct Nov Dec
Expected diarrhea cases at community level
Disruption ratio

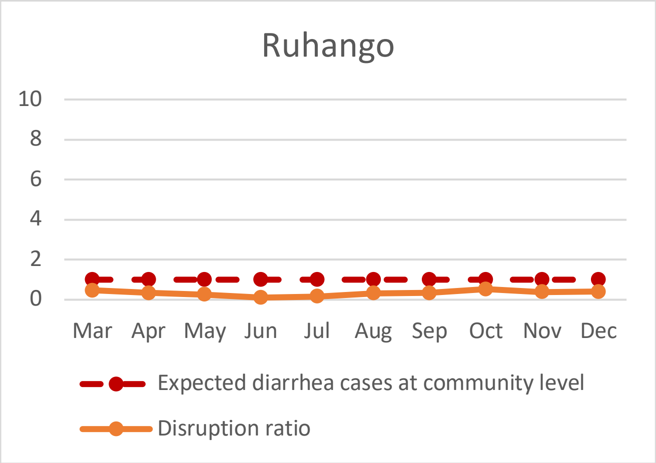
Ruhango
10
8
6
4
2
0
Mar Apr May Jun Jul Aug Sep Oct Nov Dec
Expected diarrhea cases at community level
Disruption ratio

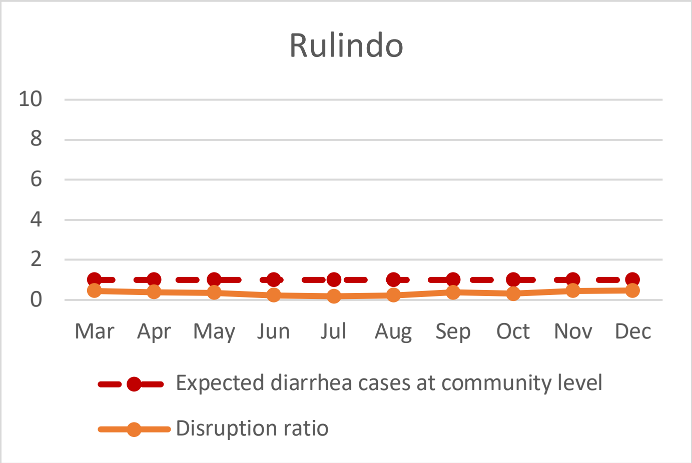
Rulindo
10
8
6
4
2
0
Mar Apr May Jun Jul Aug Sep Oct Nov Dec
Expected diarrhea cases at community level
Disruption ratio

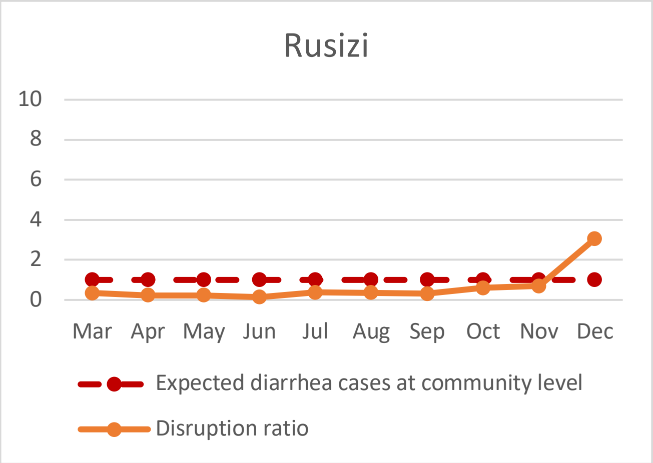
Rusizi
10
8
6
4
2
0
Mar Apr May Jun Jul Aug Sep Oct Nov Dec
Expected diarrhea cases at community level
Disruption ratio

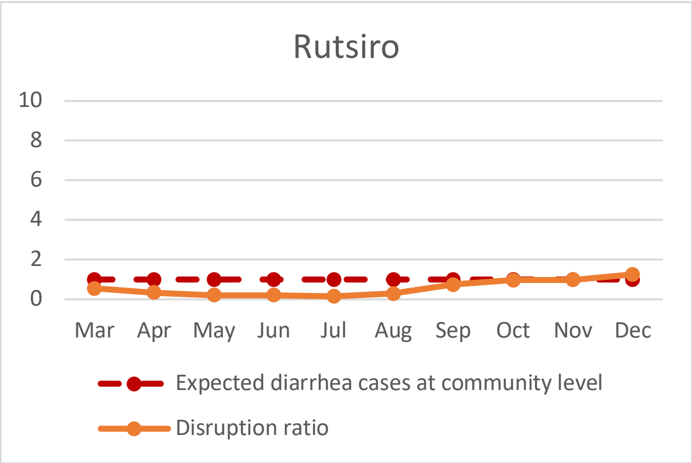
Rutsiro
10
8
6
4
2
0
Mar Apr May Jun Jul Aug Sep Oct Nov Dec
Expected diarrhea cases at community level
Disruption ratio

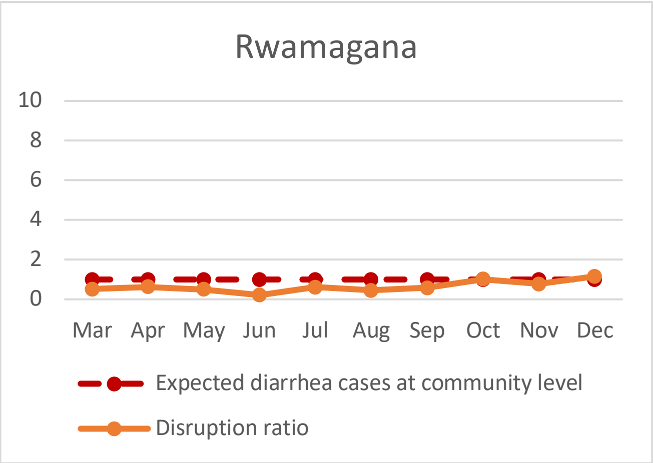
Rwamagana
10
8
6
4
2
0
Mar Apr May Jun Jul Aug Sep Oct Nov Dec
Expected diarrhea cases at community level
Disruption ratio
